# Loss-of-function of RNA-binding protein PRRC2B causes translational defects and cardiovascular malformation

**DOI:** 10.1101/2024.09.26.24313895

**Authors:** Debojyoti Das, Aleksandr V. Ivanov, Eng-Soon Khor, Feng Jiang, Liuqing Yang, Shen Zhou, Jiali He, Yui Kawakami, Lindsey Wainwright, Joshua Geiger, Fanju W. Meng, Huan Liu, Zhenggen Jin, George A. Porter, Patrick J. Murphy, Peng Yao

**Affiliations:** Aab Cardiovascular Research Institute, Department of Medicine, University of Rochester School of Medicine & Dentistry, Rochester, NY 14642; Department of Biochemistry & Biophysics, University of Rochester School of Medicine & Dentistry, Rochester, NY 14642; Department of Vascular Surgery, University of Rochester School of Medicine & Dentistry, Rochester, NY 14642; Department of Biological Sciences, University of North Texas, Denton, TX 76201; Department of Pediatrics, Medicine, and Pharmacology and Physiology, University of Rochester School of Medicine & Dentistry, Rochester, NY 14642; Department of Molecular Biology and Genetics, Cornell University, Ithaca, NY 14853; The Center for RNA Biology, University of Rochester School of Medicine & Dentistry, Rochester, NY 14642; The Center for Biomedical Informatics, University of Rochester School of Medicine & Dentistry, Rochester, NY 14642

**Keywords:** translation, PRRC2B, RNA-binding protein, alternative splicing, cardiovascular

## Abstract

Alternative splicing of pre-mRNA generates variant protein forms from a given gene, accounting for both functional redundancy and diversification. A novel RNA-binding protein, <u>Pr</u>o-rich <u>C</u>oiled- coil Containing Protein <u>2B</u> (PRRC2B), has been shown to mediate both upstream (u)ORF-dependent and independent regulation of translation initiation, which is essential for cell cycle progression and proliferation. We identified two alternative spliced isoforms of *PRRC2B* mRNA in human and mouse hearts, as well as in multiple cell types: the full-length and the exon 16- excluded isoform ΔE16. A congenital heart disease-associated human mutation-mimicking knock-in of the equivalent variant in the mouse genome results in the depletion of full-length *Prrc2b* mRNA, but not the alternative spliced truncated form ΔE16. This mouse model does not cause any apparent structural or functional cardiac disorders. In contrast, global genetic inactivation of the *Prrc2b* gene in the mouse genome, nullifying both mRNA isoforms, caused patent ductus arteriosus (PDA) and neonatal lethality. Bulk and single-nucleus transcriptome profiling analyses of embryonic mouse hearts at E18.5 demonstrate a significant overall downregulation of multiple smooth muscle-specific genes in *Prrc2b* knockout mice, likely due to a reduced smooth muscle cell number as confirmed by flow cytometry. An integrated dual-omics analysis of proteomic and translatomic changes in *Prrc2b* null mouse E18.5 embryonic hearts, combined with polysome-seq and RNA-seq in HEK293T cells, reveals enrichment of PRRC2B-regulated target mRNAs encoding essential factors required for cell proliferation and cardiovascular development. Lentiviral-shRNA-mediated knockdown of PRRC2B in human aorta-derived smooth muscle cells (HASMCs) reduces the proliferation, migration, and contractile function-related gene expression. Overall, this study demonstrates the functional and mechanistic connections between PRRC2B- mediated translational control and congenital cardiovascular development and disorders.

**Discovery bullet points:** 1. PRRC2B has two alternative splicing isoforms, full-length and exon 16-skipped (ΔE16) mRNAs in humans and mice, with redundant and specialized interactome and likely functions.

2. Full-length *Prrc2b* knockout mice exhibit no apparent cardiac phenotypes, whereas the double knockout of both isoforms results in patent ductus arteriosus and neonatal lethality in mice.

3. Multi-omics analyses of *Prrc2b* dual-isoform knockout mice reveal translation dysregulation of specific proteins in embryonic hearts and a decrease in smooth muscle cell abundance.

4. PRRC2B knockdown in human aorta-derived smooth muscle cells compromises cell proliferation, migration, and contractile gene expression.

## Introduction

Transcriptional regulation of gene expression controls cell differentiation, tissue patterning, and organ formation. Post-transcriptional messenger RNA (mRNA) metabolic processes, in addition to transcription, also play essential roles in maintaining the accuracy of developmental programs and organismal health (1,2). Post-transcriptional regulation modifies the coding capacity of mRNA and modulates the translation activity for protein synthesis. A key RNA processing pathway, alternative splicing of precursor mRNA, produces multiple isoforms of mature mRNAs that often encode distinct protein isoforms (3). These protein isoforms can perform redundant or diversified functions in cells, depending on their structural and functional relationships. In addition, translation regulatory factors modulate the rate of translation initiation or elongation, thereby precisely controlling the amount of protein products (4). However, the mechanisms by which the alternative splicing of translation regulatory factors affects cell function and organ development remain largely unknown.

<u>Pr</u>o-rich <u>C</u>oiled-coil Containing Protein <u>2B</u> (PRRC2B) is one of the members of the PRRC2 protein family, which is recognized for its RNA-binding activities (5–7). A previous study suggests that PRRC2B forms a complex with eIF4G2, a translation initiation factor that drives translation during mouse embryonic stem cell differentiation (8). Our recent findings demonstrate that the PRRC2B- eIF4G2 complex mediates the translational activation of a cohort of mRNAs encoding cell-cycle progression-related proteins by directly binding to specific RNA motifs (9). However, its biological function is still largely unknown. A previous study in the animal model showed that *Prrc2b* mRNA is highly expressed in rat whole brain tissues from early embryonic to neonatal stages, but is expressed at low levels in adulthood (10). Most recently, an endothelial cell-specific conditional knockout of the *Prrc2b* gene in mice enhanced hypoxia-induced vascular remodeling and rearrangement of cerebral blood flow, thereby reducing hypoxia-driven cognitive decline (11). However, it remains unclear whether PRRC2B is biologically vital for heart development.

Congenital heart diseases (CHD) are a group of abnormalities of the heart that develop during embryogenesis. Affecting approximately 1% of live births, CHD is a leading cause of mortality from congenital disabilities. CHD encompasses various developmental defects that affect the structure of the heart or blood vessels, such as abnormalities in cardiac vessel development (12,13), which can lead to impaired cardiac function (14). CHD can result from rare *de novo* genetic mutations, often classified as loss-of-function (LOF) mutations, in genes related to normal cardiovascular development, suggesting the essentiality of these genes for heart or vessel development. A more comprehensive human clinical study, reported for the first time, found that *de novo* mutations have significantly contributed to CHD (13). Interestingly, *PRRC2B* was one of the 12 risk genes not previously identified in CHD (13). PRRC2B has two heterozygous mutations in CHD patients, including a nonsense mutation p.R1113X and an intronic mutation (c.6381+4dupA). These two mutations are associated with mitral valve regurgitation and stenosis, and with pulmonary vein atresia and stenosis, respectively. However, the causal relationship between genotype and phenotype has not yet been established.

While previous human studies have suggested that PRRC2B may be associated with CHD, no reports are available regarding its biological role in the heart *in vivo*. This study identified two alternative spliced isoforms of the *PRRC2B* mRNA in humans and mice. We performed a bioinformatic analysis of PRRC2B in a cohort of CHD patients with cardiac genetic mutations. We then used two genetic knockout mouse models (*Prrc2b*^KI^ and *Prrc2b*^tm1b^) to investigate the role of PRRC2B in the heart from embryonic stages through adulthood. Our data suggest that the loss of both isoforms of PRRC2B causes a developmental deficiency in ductus arteriosus closure immediately after birth, whereas the full-length isoform is not required for normal embryonic cardiac development or a healthy status at the early postnatal stage. Our work reveals that PRRC2B alternative splicing generates a truncated isoform that can protect organisms against the loss of function of the full-length protein, thereby maintaining cardiovascular integrity and organismal well-being.

## Results

### Alternative splicing generates two conserved mRNA isoforms of PRRC2B in humans and mice

Recent reports from our laboratory and others have demonstrated the translation-regulatory function of PRRC2B in human cells (9,15). *PRRC2B* is highly expressed in the aorta and coronary arteries of the human cardiovascular system **(Figure S1A)**. Whereas *Prrc2b* mRNA is expressed in murine hearts as early as E12.5 and reduces moderately after birth with time into adulthood **(Figure S1B)**. We searched the NCBI and GTEx Portal databases and identified two alternative splicing (AS) mRNA isoforms of PRRC2B: a full-length (FL; 32 exons in humans and 31 exons in mice) and an exon 16-excluded (ΔE16; with no exon 16). These isoforms are evolutionarily conserved in humans and mice (**Figure 1A**). Exon 16 (2082 bp) encodes amino acids 775-1468, which contain RNA-binding Arg-Gly (RG) repeat motifs that interact with target mRNAs in humans (9), suggesting potentially reduced RNA-binding and translation activity of ΔE16 compared to the full-length protein. Consistently, our prior findings show that the 750-1500 aa of the PRRC2B protein can primarily mediate the interaction with target mRNAs (9). These two alternatively spliced isoforms were annotated across various human organs in the GTEx Portal database (**Figure S1C**). The mouse *Prrc2b* exon 16 (2,253 bp) encodes amino acids 809-1558, which bear the conserved RNA-binding RG repeat motifs, as in humans. RNA-seq of mouse heart tissues demonstrated the existence of exon-exon junction reads across exons 15 and 16, exons 16 and 17, and exons 15 and 17 (**Figure 1B**), confirming the AS event *in vivo*. Our RT-PCR data also validated the presence of the two AS mRNA isoforms in human AC16 ventricular cardiomyocyte cells, HEK293T cells (**Figure 1C**), immortalized human cardiac fibroblasts (IHCFs), and mouse hearts (**Figure S1D, E**). The protein structure has been predicted by Alpha-Fold (16,17) for human FL PRRC2B and mouse and human ΔE16 PRRC2B, but not for mouse FL proteins **(Figure 1D)**. Most regions of both protein isoforms are predicted to be intrinsically disordered regions (IDRs) (18,19), including the exon 16-encoded region. Convincingly, we found direct proteomic evidence for the existence of human ΔE16 as a peptide spanning exons 15 and 17, which was identified as <u>SSDTLAMDMRV</u>*RSPDEALPGGLSGCSSGSGHSPYALE* <u>(underlined</u> for exon 15 and *italicized* for exon 17) in the ProteomicsDB database.

**Figure 1.**
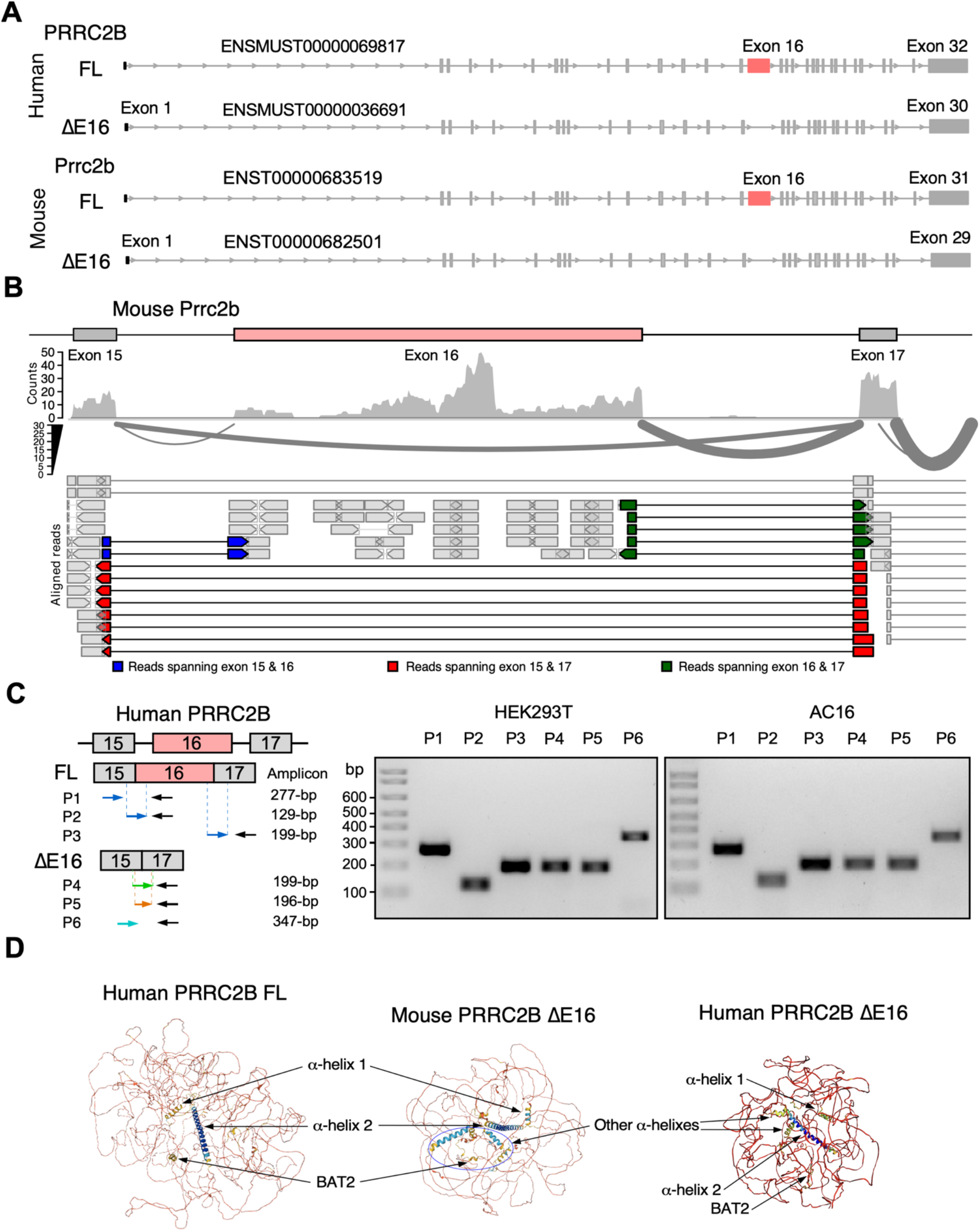
Alternative splicing isoforms of *PRRC2B* mRNA in humans and mice. **A.** Schematic of two alternative splicing isoforms of *PRRC2B* mRNA in humans and mice. **B**. RNA-seq reads mapped to exon-exon junction regions at the exons 15-17. Adult hearts from male C57BL/6J WT mice were used for RNA-seq. **C**. RT-PCR validation of alternative splicing isoforms of *PRRC2B* mRNA in HEK293T and AC16 cells. P1-6, primers 1-6. **D**. Predicted protein structure of full-length and ΔE16 PRRC2B in humans and mice by Alpha-Fold.

### Genetic variants of PRRC2B are associated with congenital heart defects in humans

Recent reports suggest that two human PRRC2B *de novo* mutations are associated with CHD characterized by vascular defects in the mitral valves or pulmonary veins (13,20). To further explore this genetic correlation, we performed gene-based burden testing of rare variants in the PRRC2B gene in 3,740 CHD probands from the Pediatric Cardiac Genetics Consortium (PCGC) (21). The test variants have a minor allele frequency (MAF) less than 0.1% in the gnomAD database and a combined annotation-dependent depletion (CADD) score greater than 20, indicating strong deleteriousness of single-nucleotide variants in the human genome. The outcome revealed significant enrichment in probands with atrial septal defects (**Table S1**; *P* < 0.0001). Hypoplastic left heart syndrome (HLHS) showed a trend toward enrichment for rare variants in *PRRC2B* (*P* = 0.02508). However, this phenotype did not remain significant after correction for multiple testing.

PRRC2B contains numerous arginine (Arg) amino acid residue sites. This amino acid is often found in RNA-binding regions due to its positive charge at physiological pH (22). Signals that indirectly assess changes in the RNA-binding capacity of PRRC2B have been tested, based on the hypothesis that structural alterations in the RNA-binding domains within the gene may contribute to congenital heart disease phenotypes. Given this, rare variants at Arg sites throughout PRRC2B were assessed for enrichment in CHD phenotypes. We observed enrichment of rare damaging variants within arginine sites in *PRRC2B* in probands with atrial septal defects compared to all other probands. Interestingly, one proband was identified as heterozygous for a *de novo* Arg-to-stop codon mutation, p.R1113X, in our analysis of the PCGC database (21). This proband had mitral regurgitation and mitral stenosis and has been previously reported as a CHD-associated gene (13). Taken together, these findings reveal a significant association between PRRC2B genetic variants and congenital heart defects in humans, implying a potential role for PRRC2B in heart development.

### Homozygous human mutation mimicry p.R1128X knock-in mice do not manifest cardiac functional defects at baseline or under stresses

The R1113X premature termination codon resides in exon 16 of the human *PRRC2B* gene. This Arg residue is highly conserved across multiple vertebrate species (**Figure S2A**). In mice, R1128 corresponds to R1113 in humans. Theoretically, the genetic mutation of R1113X in humans is supposed to cause nonsense-mediated mRNA decay to the full-length *Prrc2b* mRNA containing exon 16, but not the alternatively spliced truncated isoform without exon 16. To investigate whether the human heterozygous p.R1113X mutation contributes to any potential congenital heart defects *in vivo*, we generated a *Prrc2b* global knock-in (KI) mouse model using the CRISPR (clustered regularly interspaced short palindromic repeats) technology. The global KI mice were created by introducing an Arg-to-stop codon at the position of 1128 amino acid (p.R1128X) and two additional wobble base mutations (GA<u>C</u>TT<u>C</u>-to-GA<u>T</u>TT<u>T</u>) using a guide RNA and a homology-directed recombination DNA template (**Figure 2A**). TA cloning and DNA Sanger sequencing confirmed the successful replacement of a premature stop codon at the Arg codon into the *Prrc2b* genomic locus (**Figure 2B**). Both heterozygous and homozygous *Prrc2b* KI mice were viable, fertile, and their offspring exhibited Mendelian inheritance patterns.

**Figure 2.**
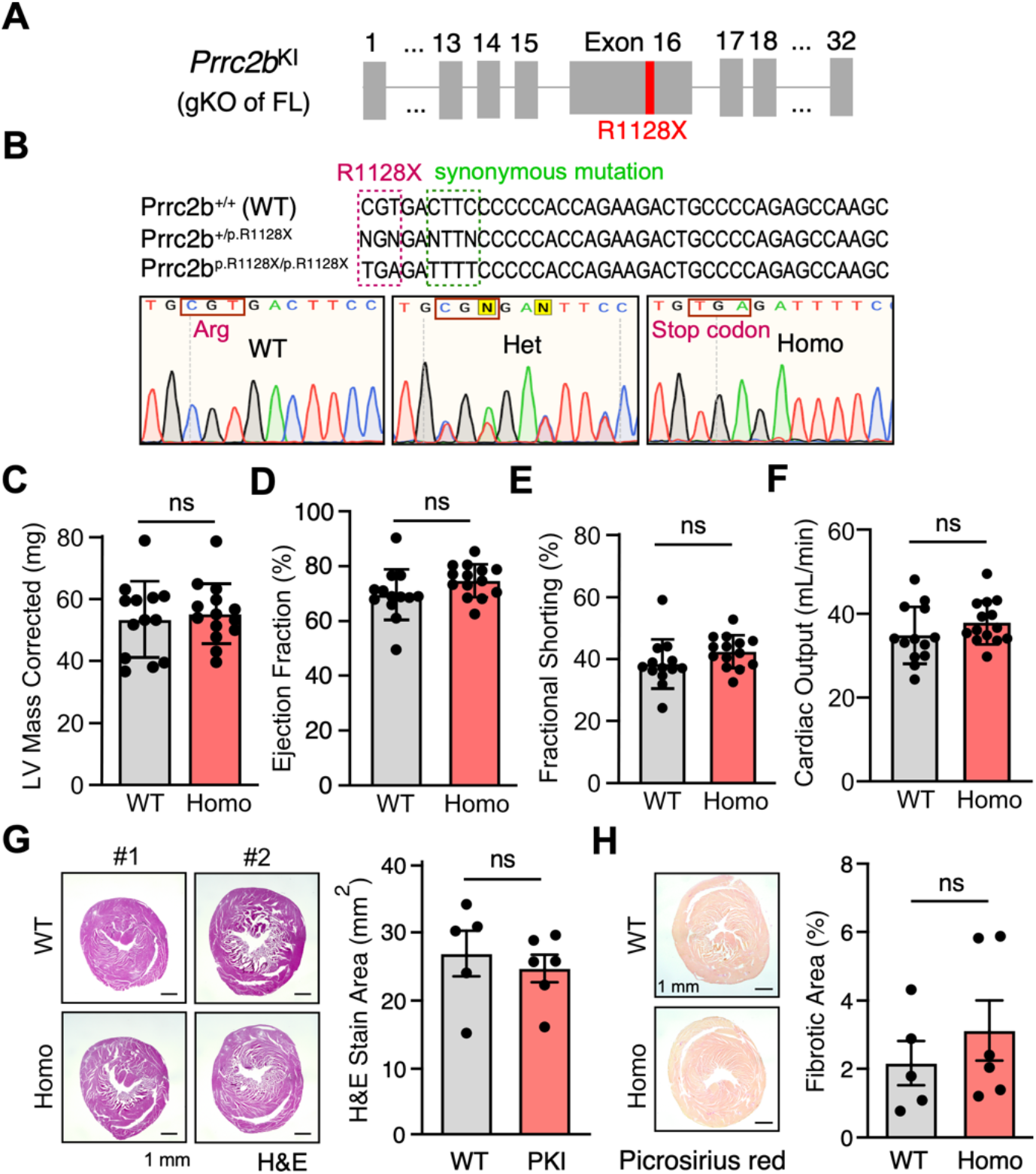
*Prrc2b*^R1128X/R1128X^ knock-in (FL-*Prrc2b*^gKO^) mice carrying a human premature termination codon mutation show inactivation of full-length *Prrc2b* mRNA without apparent cardiac phenotypes. A. Schematic of *Prrc2b*^R1128X/R1128X^ (FL-*Prrc2b*^gKO^) mouse model. **B**. Sanger sequencing confirms the genotypes of the *Prrc2b*^R1128X/R1128X^ (FL-*Prrc2b*^gKO^) mouse model. **C-F**. LV mass, ejection fraction (EF), fractional shortening (FS), and cardiac output showed no significant difference between WT control and homozygous FL-*Prrc2b*^gKO^ 3 months after birth. WT: N = 5M + 8F; KO: N = 7M + 7F. **G**. H&E images of WT and KI mice hearts at 5 months old. Heart size was quantified using Image J. WT: N = 3M + 2F; KO: N = 3M + 3F. **H**. Picrosirius red staining of WT and KI mice hearts at 5 months old. The quantification of the fibrotic area for comparison was performed using Image J. WT: N = 3M + 2F; KO: N = 3M + 3F. Data are represented as mean ± SD. An unpaired two-tailed Student t-test was performed to compare two groups for C-H. ns: not significant; * *P* < 0.05.

We hypothesized that potential ΔE16 isoform expression might compensate for the loss of full-length (FL) PRRC2B protein isoform to maintain the viability of the KI mice. As expected, the FL *Prrc2b* mRNA expression was reduced drastically by nonsense-mediated mRNA decay in the heart of *Prrc2b* p.R1128X KI (FL-*Prrc2b*^gKO^) mice, based on quantitative measurements of *Prrc2b* expression using RT-qPCR for heart tissue lysates (**Figure S2B**) (primers #1, #3, and #4). In contrast, the ΔE16 *Prrc2b* mRNA isoform expression was unaffected (primers #2, #5, and #6). To examine whether KO of FL *Prrc2b* mRNA expression causes any cardiac functional changes, echocardiography was performed to measure the left ventricular (LV) function. We observed no significant changes in LV mass in the KI mice up to 3 months after birth (**Figure 2C; Table S2**). When comparing WT and KI mice, ejection fraction, fractional shortening, or cardiac output did not change from 1 to 3 months post-birth (**Figure 2D-F; Table S2**). After 5 months, no significant differences in heart size or collagen deposition between KI and WT mice were detected (**Figure 2G, H**). H&E imaging analysis did not reveal any apparent structural differences in multiple organs of KI versus WT mice, including the brain, kidney, liver, lung, skeletal muscle (**Figure S2C**), spleen, and thymus (data not shown). These results suggest that the loss of full-length PRRC2B in mice does not cause significant cardiac pathological phenotypes, such as cardiac hypertrophy, fibrosis, or LV functional decline, at baseline in adulthood. In addition, to explore whether KI mice are more susceptible to cardiac stress than WT mice, we subjected both groups of mice to minipump- mediated infusion of combined angiotensin II (Ang II) and epinephrine (PE) as neurohumoral stimuli, compared with vehicle. We did not observe significant differences between WT and KI mice in heart morphology, LV mass, heart rate, ejection fraction, stroke volume, cardiac output, and LV geometric dimensions at the systolic or diastolic phase (**Figure S2D-K**).

### Homozygous *Prrc2b*^tm1b-/-^ mice exhibit patent ductus arteriosus and neonatal lethality

Our phenotyping data of *Prrc2b* p.R1128X KI (FL-*Prrc2b*^gKO^) mice suggested that loss of the FL PRRC2B isoform does not significantly affect cardiac structure or function (**Figure 2**). This suggests that the ΔE16 PRRC2B isoform may compensate for the loss of the full-length protein. Therefore, we generated a mouse model with both alternative-spliced *Prrc2b* isoforms nullified, termed *Prrc2b*^tm1b-/-^. The tm1b-LacZ tagged null allele was generated by deletion of the critical fourth exon of the *Prrc2b* genomic locus and the neomycin cassette using a constitutive CMV-Cre recombinase that recognizes loxP sites **(Figure 3A)**. This allele is considered an authentic knockout, as skipping over the LacZ cassette cannot restore *Prrc2b* expression.

**Figure 3.**
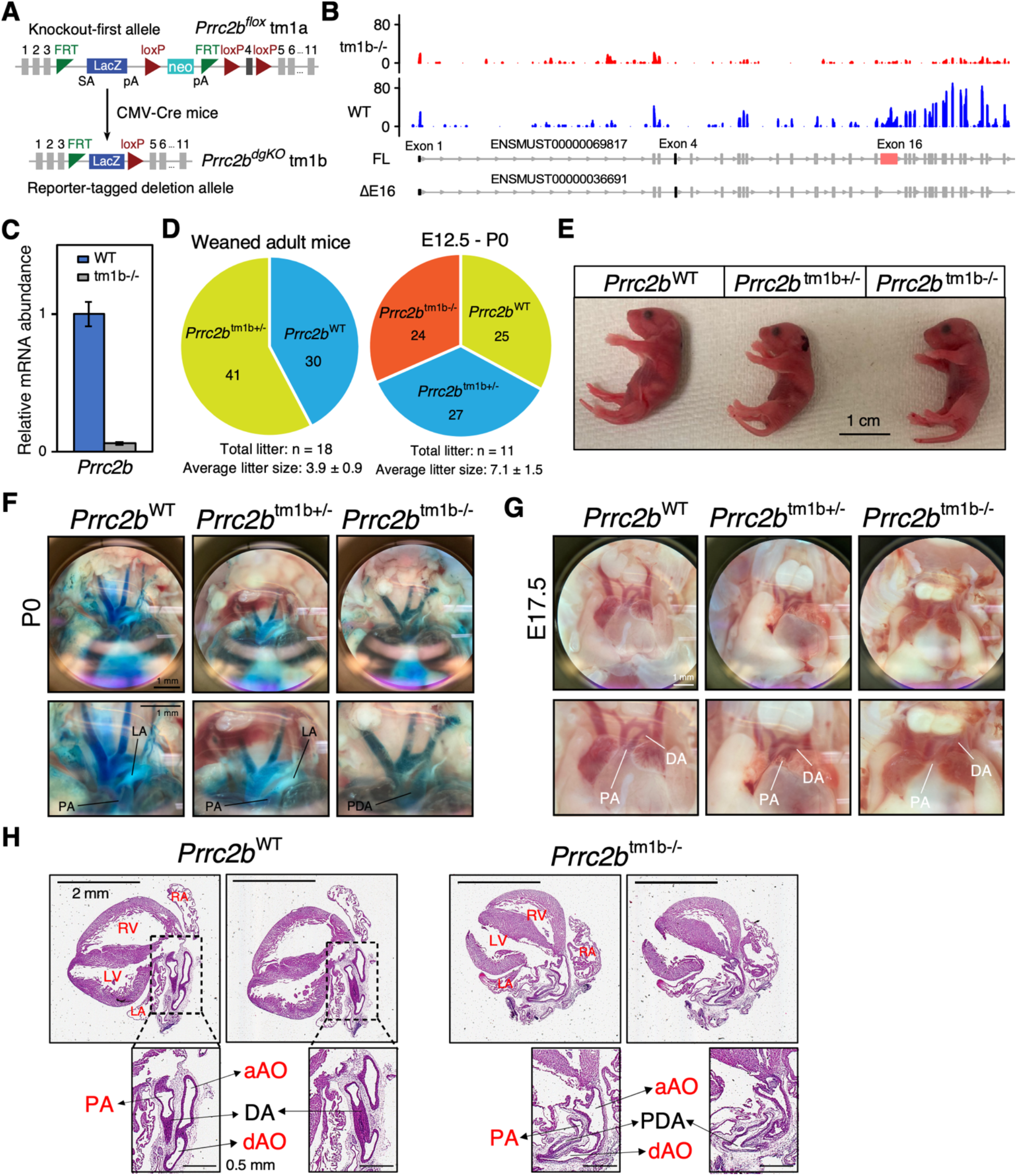
Mice lacking two *Prrc2b* alternative splicing isoforms manifest patent ductus arteriosus (PDA) and die perinatally. **A.** Schematic representation of the generation of the *Prrc2b* global KO mouse model (*Prrc2b*^tm1b-/-^) by crossing *Prrc2b*^tm1a^ with CMV-Cre mice. Both full-length and ΔE16 *Prrc2b* isoforms are knocked out. **B**. Integrative Genomics Viewer (IGV) plot of RNA-seq measurement of *Prrc2b* mRNA expression across all the exons. **C**. RT-qPCR analysis of *Prrc2b* mRNA expression in whole hearts of WT and *Prrc2b*^tm1b-/-^ mice. **D**. Number of weaned adult *Prrc2b*^tm1b-/-^ mice and prenatal *Prrc2b*^tm1b-/-^ mice (E12.5 – E18.5) during heterozygous breeding, indicated by their respective pie charts. **E**. Images of mice at P0. *Prrc2b*^-/-^ pups were found dead as early as 7 hours postnatally, while *Prrc2b*^+/-^ remained alive. Scale bar: 1 cm. **F**. Images of hearts with Coomassie blue dye injection from WT, *Prrc2b*^+/-^, and *Prrc2b*^-/-^ at P0. Scale bar: 1 mm. PA, pulmonary artery; DA, ductus arteriosus; LA, ligamentum arteriosum; PDA, patent ductus arteriosus. **G**. Images of hearts in the bright field from WT, *Prrc2b*^+/-^, and *Prrc2b*^-/-^ at E17.5. Scale bar: 1 mm. **H**. H&E images of WT and homozygous tm1b KO hearts at P0. LV: left ventricle, RV: Right ventricle, LA: left atrium, RA: right atrium, aAO: ascending aorta, dAO: descending aorta, DA: ductus arteriosus, PA: pulmonary artery, PDA: patent ductus arteriosus.

The elimination of the fourth exon at the genomic level was validated by DNA genotyping **(Figure S3A)**. In addition, the absence of both FL and ΔE16 *Prrc2b* mRNA transcripts in *Prrc2b*^tm1b-/-^ mice was confirmed by RNA-seq and agarose gel electrophoresis following real-time PCR **(Figure 3B, C, S3B, C)**. This suggests that the phenotypes observed in *Prrc2b*^tm1b-/-^ mice result from the loss of function of both *Prrc2b* AS isoforms. We observed preweaning lethality in homozygous *Prrc2b*^tm1b-/-^ mice across 18 litters from heterozygote breeding **(Figure 3D, left)**. This suggested that *Prrc2b*^tm1b-/-^ pups died either *in utero* or perinatally after a normal vaginal delivery. Consistent with this, we have obtained 24 viable *Prrc2b*^tm1b-/-^ pups in 11 litters ranging from different prenatal stages (E12.5, E13.5, E14.5, E17.5, and E18.5) to the early postnatal stage (P0) **(Figure 3D, right)**. This suggests that *Prrc2b*^tm1b-/-^ mice survive to the prenatal stage. Subsequently, we successfully captured the viable *Prrc2b*^tm1b-/-^ neonatal mice as early as 7 hours post-birth **(Figure 3E)**. The LacZ cassette was expressed in tissues where the Prrc2b gene was knocked out. β-Galactosidase staining was used to examine the tissue expression of *Prrc2b* at different developmental stages and across multiple organs. A dark blue LacZ signal was evident in *Prrc2b*^tm1b-/-^ and *Prrc2b*^tm1b+/-^ but not in WT embryonic hearts isolated at E12.5 **(Figure S3D)**. The LacZ signals were also detected in the heart, brain, pituitary, aorta, olfactory bulbs, bladder, and thymus. Still, they were barely detected in the skeletal muscle and lung from the *Prrc2b*^tm1b+/-^ and WT adult mice (4-month-old) **(Figure S3E)**. This suggests that the *Prrc2b* gene promoter is actively transcribed across *multiple or*gans and cell types, spanning from prenatal to late postnatal stages.

As *PRRC2B* human mutations have been reported to be associated with mitral valve stenosis or pulmonary vein atresia and stenosis (13,20), we next examined the state of the blood vessels in the heart. We injected Coomassie blue dye into the LV of isolated hearts from WT control and *Prrc2b*^tm1b-/-^ mice. Interestingly, we observed that the ligamentum arteriosum (LA) has a white band with the exclusion of the blue dye in the *Prrc2b*^tm1b-/-^ heart but not in the WT heart, suggesting the failure of ductus arteriosus (DA) closure at P0 **(Figure 3F)**, leading to patent ductus arteriosus (PDA). In contrast, the PDA did not occur at the earlier time point (E17.5) in the live *Prrc2b*^tm1b-/-^ prenatal mice **(Figure 3G)**. At E17.5, DA shunts blood from the PA to the aorta in the embryonic/fetal circulation. There is no difference among the three genotypes (WT, *Prrc2b*^tm1b+/-^, and *Prrc2b*^tm1b-/-^) at E17.5, as blood is present throughout the DA vessel. However, at P0, the transformation of DA into LA, which marked the closure, occurred in *Prrc2b*^tm1b+/-^ and *Prrc2b*^WT^, whereas PDA was observed in *Prrc2b*^tm1b-/-^ as the DA remained open. This phenotype was not seen in heterozygous or WT littermates **(Figure 3F)**. Hematoxylin and eosin (H&E) staining of the frontal section of the heart tissues of *Prrc2b*^tm1b+/-^ and WT mice further confirmed the sustained opening of the PDA in the homozygous KO hearts but not in the control hearts **(Figure 3H)**. Together, these findings suggest that *Prrc2b* loss-of-function leads to PDA in mice, resulting in neonatal lethality within approximately 24 hours after birth, consistent with the known consequences of persistent PDA that often causes neonatal lethality in mice (23,24). Also, comparing the phenotypes between *Prrc2b* p.R1128X KI and *Prrc2b*^tm1b-/-^ mice, we indicate that the ΔE16 PRRC2B protein isoform may compensate for the loss of full-length PRRC2B function *in vivo*.

### Knockdown of PRRC2B leads to abnormal cardiac morphology in zebrafish

To determine whether PRRC2B is evolutionarily crucial and conserved in normal cardiac development, we knocked down *Prrc2b* gene expression in zebrafish using both splice-blocking and translation-blocking morpholino (MO) antisense oligonucleotides (**Figure S3F**). RT-PCR data suggest that intron 3 (106 bp) of *Prrc2b* was retained in the mRNA, which yielded a higher band as observed in the agarose gel (**Figure S3G**). As a result, introducing three premature stop codons within this intron is likely to trigger nonsense-mediated mRNA decay and thus reduce the protein expression of PRRC2B. We have observed pericardial edema and heart looping defect present only in five *Prrc2b* morphant zebrafish (5/30; penetrance of 16.7%) injected with splice-blocking MO but not in control zebrafish (0/25) **(Figure S3H)**. However, 86.7% of the *Prrc2b* morphant zebrafish (52/60) injected with translation-blocking MO were found dead 3 days post-injection, likely due to severe abnormalities in the heart development, as a low amount of PRRC2B protein can be produced in this case. The eight surviving fish with translation-blocking MO also showed pericardial edema, while none of the morphant zebrafish injected with control MO showed any cardiac phenotypes (0/18) (**Figure S3I**). However, because no suitable antibody is available for the zebrafish PRRC2B protein, it is challenging to determine PRRC2B protein expression levels in *Prrc2b* morphant zebrafish injected with a translation-blocking MO. This data suggests the importance of PRRC2B in maintaining normal cardiac development in zebrafish, as early as 36 hours post-fertilization (hpf).

### Transcriptomic profiling reveals alterations of gene expression related to translation, mitochondria, smooth muscle cells, and cardiovascular development in *Prrc2b*^tm1b-/-^ mice

PRRC2B has been identified as an RNA-binding protein by unbiased mRNA interactome capture analysis using mass spectrometry (5–7). Our recent findings suggest that PRRC2B can bind specific RNA motifs and activate translation of a selective cohort of cell proliferation-related mRNAs (9). Another study shows that PRRC2B promotes leaky scanning of uORFs, thereby enhancing translation of main ORFs (15). To shed light on the molecular function of PRRC2B in the heart, we performed paired-end RNA-seq on whole hearts of *Prrc2b*^tm1b-/-^ and WT mice at E18.5. We identified 78 genes that were significantly dysregulated in knockout (KO) hearts compared to WT controls (*P*_adj_ < 0.05, without a Log_2_ FC cutoff) **(Figure 4A, B, S4A, Table S3)**. Of these, 42 were upregulated, and 36 were downregulated. After adopting a highly stringent criterion (*P*_adj_ < 0.05 and |Log_2_ FC| > 1), only 12 dysregulated genes were identified, including 10 upregulated and 2 downregulated genes **(Figure S4B, Table S3)**. Using the web tools of database for annotation, visualization, and integrated discovery (DAVID), gene ontology analysis of 78 differentially expressed genes (*P*_adj_ < 0.05, without a Log_2_ FC cutoff) revealed that pathways related to translation and mitochondria, such as cytoplasmic translation, mitochondrial respiratory chain complex I assembly, and mitochondrial ATP synthesis coupled to proton transport, were among the top downregulated biological processes in KO hearts **(Figure 4C, S4C)**. Critical genes in the translation machinery and mitochondria were significantly reduced at the mRNA steady-state levels, including ribosomal protein genes (*Rps29*, *Rps28*, *Rpl37*, *Rpl39*), translation factors (*Eif4g1*, *Eif3l*, *Eif4h*, *Eif5a*), essential mitochondrial genes (*Lars2*, *Rars2*, *Tomm7*, *Ndufb4*, *Uqcr11*, *Ndufa1*, *mt-Nd2*, *mt-Atp6*, and *mt-Cytb*), and most importantly, cell cycle-related genes (*Foxm1*, *Aurka*, *Ccnd1*, *Ccna2*, and others), as well as genes involved in regulation of muscle cell contraction (*Acta2*, *Tmod1*, *Lmod1*). In addition, multiple pathways related to cardiovascular development and blood vessel formation were among the top upregulated biological processes in KO hearts, including extracellular matrix (ECM) organization (*Meox1*, *Thbs1*, *Postn*, *Mmp14*, *Abi3bp*, *Col11a1*, *Adamtsl2*, *Ptx3*, *Matn4*), negative regulation of smooth muscle cell proliferation (*Bmp2*, *Igfbp5*, *Apoe*), and blood vessel remodeling (*Sema3c*, *Rspo3*, *Igf1*) **(Figure 4D, S4C)**. The small number of differentially expressed genes that are significantly altered upon *Prrc2b* knockout aligns well with PRRC2B’s function in regulating translation initiation of a selective cohort of mRNAs (9). Interestingly, a key translation initiation factor, *Eif1* (eukaryotic translation initiation factor 1), is also significantly increased as a potential compensatory effect upon the loss of *Prrc2b* (**Figure 4A**).

**Figure 4.**
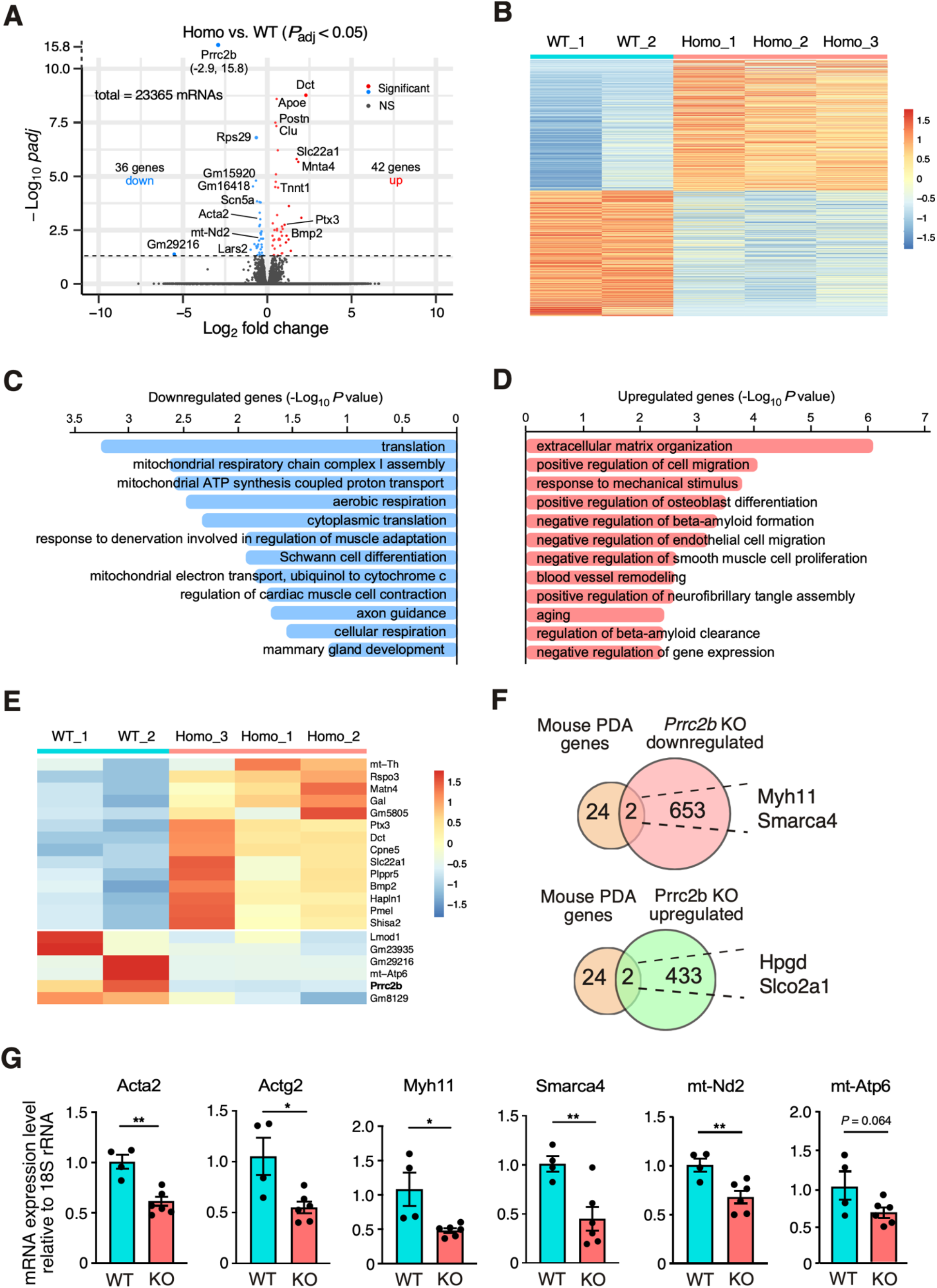
Bulk RNA-seq analysis of *Prrc2b* global knockout hearts shows reduced smooth muscle cell contraction and mitochondrial respiration-related gene expression. A. Volcano plot of RNA-seq of WT and *Prrc2b*^tm1b-/-^ hearts at E18.5. Log_2_ Fold Change (*Prrc2b*^tm1b-/-^ / WT) is plotted as the X-axis, while -Log10 *P*-values are plotted as the Y-axis. Genes with |Log2 Fold Change| > 1 and *P*-value < 0.05 are colored red or blue. WT: N = 2; *Prrc2b*^tm1b-/-^: N = 3. **B**. Heatmap of RNA-seq of WT and *Prrc2b*^tm1b-/-^ hearts. TPM (transcript per million) is plotted. Values are scaled for each row. **C and D**. Gene ontology analysis of upregulated and downregulated genes in RNA-seq of WT and *Prrc2b*^tm1b-/-^ hearts. **E**. A zoomed heatmap showing the top 20 dysregulated genes in (**B**). TPM is plotted. Values are scaled for each row. **F**. Venn diagrams showing the overlaps between PDA genes and dysregulated genes in *Prrc2b*^tm1b-/-^ hearts. Genes with |Log2 Fold Change| > 1 and *P* < 0.05 are considered dysregulated. **G**. RT-qPCR validation of dysregulated genes related to smooth muscle cell contraction and mitochondrial respiratory chain complex in *Prrc2b*^tm1b-/-^ hearts. 18S rRNA is used as the normalizer. Values are plotted as relative values to WT hearts. Data is shown as mean ± SD. Technical replicates for each biological sample are plotted (WT: N = 2; *Prrc2b*^tm1b-/-^: N = 3). An unpaired two-tailed Student t-test was performed to compare two groups for G. * *P* < 0.05; ** *P* < 0.01.

Among the top 20 differentially expressed genes (DEGs), we observed that *Lmod1* (Leiomodin 1, an essential protein for assembly and contractility of the actin cytoskeleton in smooth muscle cells) and *mt-Atp6* (a key mitochondrial-encoded subunit of ATP synthase) were significantly reduced (*P* < 0.05). In contrast, multiple transmembrane or secretory protein-encoding genes *Matn4*, *Ptx3*, *Slc22a1*, *Plppr5*, *Dct*, *Cpne5*, *Pmel*, *Shisa2*, and *Bmp2* were significantly increased (*P* < 0.05) **(Figure 4E)**. The RNA-seq reads of *Prrc2b* mRNA were drastically decreased in *Prrc2b* null hearts, confirming the genetic KO effect at the gene expression level. Intriguingly, overlapping our identified DEG with the reported list of genes involved in mouse or human PDA revealed four mouse PDA genes **(Figure 4F)**, including *Myh11* and *Smarca4*, which were reduced in *Prrc2b* KO heart samples. qPCR validation further confirmed the significantly reduced expression of *Myh11* and *Smarca4* mRNAs, as well as other SMC mRNAs, including *Acta2* and *Actg2*, and mitochondrial mRNAs such as *Nd2* and *Atp6* **(Figure 4G)**, in E18.5 KO hearts. This suggests that the vascular smooth muscle contraction essential for ductus arteriosus closure may be impaired in the *Prrc2b* knockout heart, which contributes to the development of PDA.

### Single-nucleus analysis of transcriptome in wild-type and *Prrc2b*^tm1b-/-^ E18.5 hearts indicates a reduced number of smooth muscle cells

PRRC2B is an RNA-binding protein expressed in multiple organs and cell types. To further examine the cell type-specific gene expression in KO hearts, we conducted single-nucleus RNA-sequencing (snRNA-seq) to measure gene transcription changes across distinct cardiac cell types in WT and homozygous *Prrc2b*^tm1b-/-^ mice (**Figure 5A**). We obtained expression profiles from 13,665 nuclei, with a median of 3,433 RNA-seq reads per nucleus. These include 4,660 nuclei from WT mouse hearts, 4,413 from *Prrc2b*^tm1b+/-^ hearts, and 4,592 from *Prrc2b*^tm1b-/-^ hearts.

**Figure 5.**
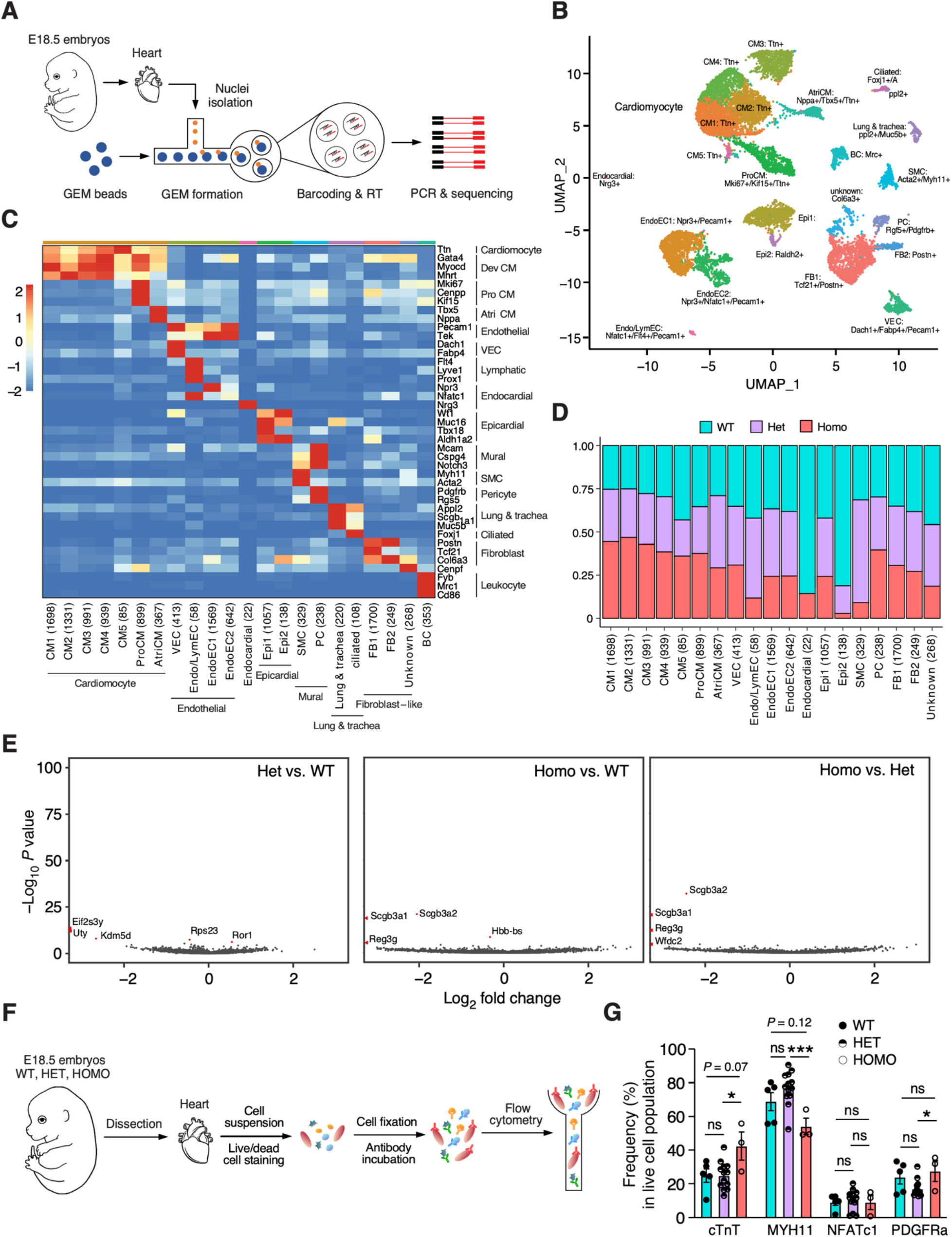
Single-nucleus (sn)RNA-seq analysis of *Prrc2b* global knockout hearts. **A**. A sketch showing the workflow of snRNA-seq for homozygous *Prrc2b*^tm1b-/-^ gKO, heterozygous *Prrc2b*^tm1b+/-^, and WT control hearts at E18.5. N = 3 hearts of each individual genotype in the same litter or two litters around the same time of birth were combined for the snRNA-seq analysis. **B**. A UMAP (uniform manifold approximation and projection) presentation of clustering results based on the top 3000 variant features. 19 clusters were identified and labeled with cell type and top marker genes. **C**. Heatmap showing the expression of well-established marker genes in each cluster. Scaled and normalized expressions were plotted. **D**. A grouped bar plot showing the relative abundance of WT, Het, and Homo cells in each cell type. The Y-axis represents the fraction of total cells in each cell type. **E**. Volcano plots showing the DEGs in smooth muscle cells. Genes with significantly differential expression (Log_2_ FC > 0.2 and Bonferroni correction adjusted *P* < 0.05) were red-colored. Ven.CM, ventricular cardiomyocytes; Atri.CM, atrial cardiomyocytes; FB, fibroblasts; EC, endothelial cells; SMC, smooth muscle cells; BC, blood cells; VEC, vascular EC; Epi, epicardial cells; PC, pericytes; ProCM, proliferating CM; Endo, endocardial; Lym, lymphatic. **F, G**. Flow cytometry-based quantification of cell numbers in freshly extracted E18.5 hearts from WT (N = 5), heterozygous (N = 16), and homozygous (N = 3) *Prrc2b* KO mice using cell type-specific antibodies. Antibodies against cTnT, MYH11, NFATc1, and PDGFRα were used to detect cardiomyocytes, smooth muscle cells, epicardial cells, and cardiac fibroblasts, respectively.

Utilizing Seurat, we identified 22 distinct clusters, which were annotated based on the RNA expression patterns of well-established lineage-specific marker genes (**Figure 5B, C, S5A**). These 16 clusters collectively represent seven major cardiac cell types: ventricular cardiomyocytes (Ven.CM), atrial cardiomyocytes (Atri.CM), fibroblasts (FB), endothelial cells (EC), smooth muscle cells (SMC), pericytes, and epicardial cells. We observed a good quality of the data based on the number of genes (nfeature), number of reads (ncount), percentage of mitochondrial-coding genes (**Figure S5B-D**), and correlation between the number of genes (nfeature, y-axis) and number of reads detected in each nucleus (**Figure S5E**). Notably, we found that the relative number of nuclei or cells was significantly decreased in SMC, endocardial/lymphatic EC, epicardial cells, and endocardial cells from homozygous KO hearts compared to WT hearts (**Figure 5D**). In contrast, the abundance of CM nuclei in homozygous KO hearts increased compared to WT ones (**Figure 5D**). Subsequently, we examined the differential gene expression patterns between WT and *Prrc2b* KO hearts. Intriguingly, we did not find evidence supporting significant gene expression differences in SMC from KO hearts compared to WT hearts, suggesting a reduced SMC cell number in the KO hearts compared to the control hearts, which led to reduced SMC-related gene expression in the bulk RNA-seq analysis as shown above (**Figure 5E; Table S4**). Using cell type-specific antibodies, flow cytometry-based quantification of cell numbers in freshly extracted E18.5 hearts from WT, heterozygous, and homozygous *Prrc2b* KO mice confirmed the increase in the number of cardiomyocytes (cTnT- positive), accompanied by the decrease in the number of smooth muscle cells (MYH11-positive) and epicardial cells (NFATc1-positive), with no significant change in fibroblasts (PDGFRα-positive) (**Figure 5F, G, S5F**).

### Translatomic profiling reveals translational dysregulation of cell proliferation and cardiac development-related mRNAs

We previously demonstrated that PRRC2B plays a critical role in the translational regulation of specific target mRNAs for protein synthesis in HEK293T cells (9). To understand the translation regulatory function of PRRC2B in the heart *in vivo*, we performed polysome profiling coupled with RNA sequencing of pooled translational fractions (polysome-seq) with the E18.5 hearts from homozygous *Prrc2b*^tm1b-/-^ KO mice and WT control mice (**Figure 6A**). The overall polysome profile suggests a moderate decrease in 80S monosomes, consistent with our reported role of PRRC2B in translation initiation (9). Translation efficiency (TE) of individual mRNA was calculated as the ratio of polysome-associated mRNA reads to monosome-associated mRNA reads, normalized to total RNA abundance. We identified a specific cohort of mRNAs with significantly decreased or increased translation efficiency (**Figure 6B; Table S5**). Gene Ontology analysis suggests that mRNAs with reduced translation efficiency were enriched in fatty acid metabolism, the neuronal system, actin cytoskeleton organization, positive regulation of cell migration, hematopoietic stem cell proliferation, negative regulation of apoptosis, and chromatin organization (**Figure 6C**). Beyond the enriched GO pathways, functional dissection of genes involved in smooth muscle cell biology and vascular remodeling identified the top 8 genes, including HHIPL1 (promotes SMC proliferation and migration), EFEMP2 (ECM protein essential for elastic fiber formation and arterial wall stability and remodeling), ARHGEF26 (activates Rho GTPase signaling and regulates SMC contraction and synthetic phenotype transition), DIAPH3 (regulates SMC migration and cytoskeletal remodeling), VASP (mediates NO/cGMP signaling in SMC), ADCY4 (produces cAMP to regulate SMC relaxation and vascular tone), SNAI2 (controls mesenchymal transition, promotes fibroblast activation, and regulates vascular remodeling), THBS1 (regulates SMC proliferation and migration and cardiac fibrosis), and SAMD1 (influences atherogenesis in vascular remodeling). To confirm potential mRNA targets regulated by PRRC2B, we performed polysome profiling followed by RT-qPCR to determine whether *HHIPL1* and *EFEMP2* mRNAs are translationally regulated by PRRC2B. Our data suggest that *HHIPL1* and *EFEMP2* mRNAs were shifted from the heavy to the lighter polysome fractions, but not for *ACTB* mRNA (**Figure 6D**), indicating translational repression for these two mRNAs identified from the polysome-seq screen. In contrast, GO analysis also suggests that mRNAs with increased translation efficiency were enriched in protein ubiquitination, monoatomic cation transport, negative regulation of the canonical Wnt signaling pathway, positive regulation of the canonical NF-kB signal transduction, chromosome segregation, and chromatin organization (**Figure 6C**).

**Figure 6.**
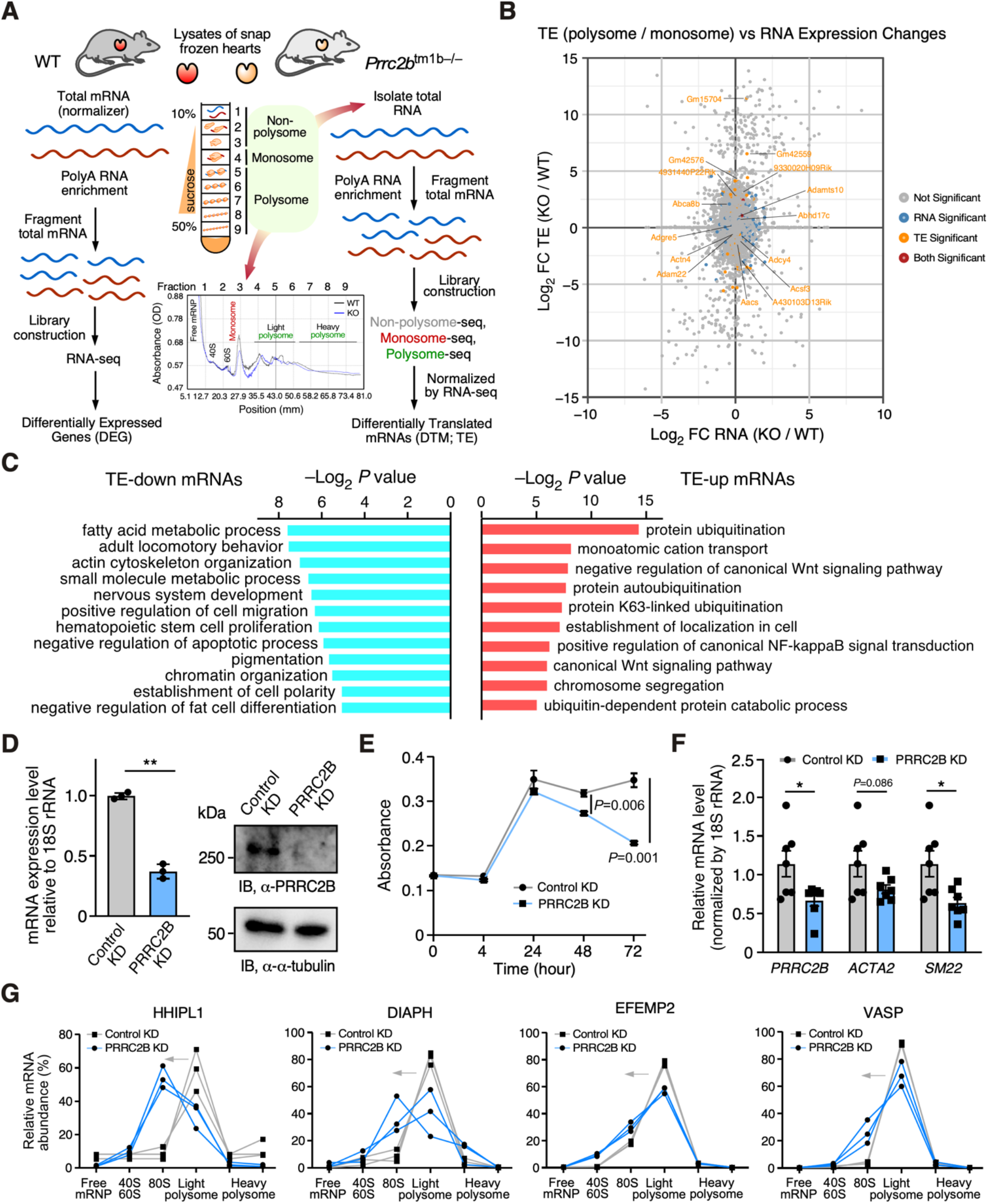
Polysome profiling-based translatomic profiling of *PRRC2B* knockout embryonic mouse hearts and functional validation in human aorta-derived smooth muscle cells. A. Schematic model of polysome profiling of E18.5 *Prrc2b*^tm1b-/-^ gKO compared with WT control hearts, followed by RNA-seq and polysome-seq. A representative polysome profile was shown for both gKO and WT hearts from male mice (N = 3). **B**. Dot plot showing differentially expressed genes (DEGs) and differentially translated mRNAs (DTMs) identified by RNA-seq and polysome- seq in *Prrc2b* gKO and WT control hearts. **C**. Gene ontology analysis of significantly downregulated or upregulated mRNAs at the translation level (*P* < 0.05). **D**. Lentiviral knockdown of *PRRC2B* in human aorta-derived smooth muscle cells (HASMCs) confirmed by RT-qPCR and Western blot. **E**. Quantitative analyses of cell proliferation rate using MTT assay in HASMCs upon *PRRC2B* knockdown. **F**. RT-qPCR analyses of *PRRC2B* and SMC marker gene expression in HASMCs upon *PRRC2B* knockdown. **G**. Polysome profiling coupled with RT-qPCR for SMC functionally related mRNAs in HASMCs upon *PRRC2B* knockdown. This experiment was repeated three times (biological replicates), and all the results were shown. Data are represented as mean ± SD. An unpaired two-tailed Student t-test was performed to compare two groups for D-F. * *P* < 0.05; ** *P* < 0.01.

### Knockdown of PRRC2B leads to reduced proliferation, migration, and contractile gene expression in human aorta-derived smooth muscle cells

*Prrc2b* global knockout mice exhibited a severe PDA phenotype, which is generally caused by compromised cell proliferation or contractile function in the vascular smooth muscle cells (VSMC). To determine if PRRC2B is required for normal smooth muscle function at the cellular level, we cultured human aorta-derived smooth muscle cells (HASMCs) and knocked down the PRRC2B gene efficiently using human PRRC2B-specific lentiviral-shRNA over 72 hours at both mRNA and protein levels (**Figure 6D**). HASMC growth was significantly reduced at 48 and 72 hours after gene knockdown (**Figure 6E**). Also, migration capacity was compromised in HASMCs upon PRRC2B knockdown (**Figure S6A**). At the endpoint, we observed a moderate decrease in SMC contractile function-related marker gene expression by 30-50% for *ACTA2* and *SM22*, respectively (**Figure 6F**), indicating the critical role of PRRC2B in regulating smooth muscle cell function.

Because the polysome-seq analysis was conducted in whole hearts from *Prrc2b* global KO mice, we next used cultured HASMCs to further validate the role of PRRC2B in translational regulation in smooth muscle cells, the major cell type involved in PDA pathogenesis. To examine the role of PRRC2B in regulating the translation of mRNAs related to SMC and blood vessel function, we performed polysome profiling in HASMCs upon 72 hours of PRRC2B knockdown and detected the location of mRNAs of *HHIPL1*, *DIAPH3*, *EFEMP2*, and *VASP*. These SMC functionally related mRNAs were shifted from translating polysome fractions to the monosome or non-translating fraction (**Figure 6F**), consistent with their translational inhibition observed in the polysome-seq analysis in E18.5 KO mouse hearts (**Figure 6B; Table S5**). In contrast, the mRNA association level with polysomes remained nearly unchanged for *ATCB* (encoding β-actin) and modestly decreased for ACTA2 (encoding α-SMA) (**Figure S6B**). These results indicate the essential function of PRRC2B in maintaining basic cellular activities in smooth muscle cells by regulating the translation of mRNAs required for SMC functional integrity.

### Quantitative mass spectrometry analysis confirms specific proteomic changes enriched in cell cycle and muscle contraction-related proteins

The efficiency of transcription and translation regulates the proteomic profile, which dictates the functional output in cells and organs. To determine whether protein expression is affected in *Prrc2b*^tm1b-/-^ mice, we subjected homozygous KO and WT control mouse hearts to mass spectrometry analysis and identified a reduction in a specific protein cohort (**Figure S6C; Table S6**). GO analysis of significantly downregulated proteins was enriched in five major pathways (*P* < 0.05 without Log_2_FC cutoff): positive regulation of G1/S transition of the mitotic cell cycle, DNA replication, protein neddylation, mitochondrial respiratory chain complex IV assembly, and translation (**Figure S6D**). In contrast, GO analysis failed to identify enriched pathways for upregulated proteins due to the limited number of genes. We further investigated downregulated proteins with a cut-off of Log_2_FC < –0.75. We revealed multiple highly enriched pathways, including mitochondrial translation, mitotic cytokinesis, membrane fission, glycolysis, epithelial cell migration, and cardiac muscle contraction (**Figure S6E**). These changes indicate a compromised expression of specific functional proteins necessary for maintaining normal cardiac cell structure and function.

### Insights into conserved PRRC2B-regulated target mRNAs across cell types and species

The PRRC2B gene is ubiquitously expressed across human cell types and organs (**Figure S1A, C**), suggesting a more generalizable role in mRNA translation regulation, maintenance of cellular function, and organismal development. To identify potential commonly shared PRRC2B-regulated target mRNAs in humans and mice across different organs or cell types, we created a CRISPR-derived *PRRC2B* KO human HEK293T (human embryonic kidney epithelial cells) cell line as confirmed by DNA Sanger sequencing and Western blotting (**Figure S7A, 7A**). In the validated *PRRC2B* KO HEK293T cell line, we confirmed by Sanger sequencing that three copies of *PRRC2B* DNA alleles were disrupted by random insertions or deletions within the exon 7 that is shared by both FL and ΔE16 PRRC2B. Therefore, both alternative spliced *PRRC2B* mRNA isoforms were depleted in the KO cells. Polysome profiling and puromycin incorporation assays indicated no apparent changes in global mRNA translation and *de novo* protein synthesis (**Figure 7B, S7B**). RNA-seq analysis uncovered changed gene expression at the mRNA steady-state level in *PRRC2B* KO human cells (**Figure 7C, S7C-E; Table S7**). ECM-related genes were significantly reduced, while neurogenesis, ion channel, and cytoskeleton-related genes were increased (**Figure 7D**). The reduction in ECM gene expression is opposite to the increase in *Prrc2b* tm1b KO hearts, suggesting that ECM remodeling in the whole heart is a secondary effect of multiple cardiac cell types rather than an autonomous effect of a single cell type. In addition, PRRC2B is primarily a translation regulatory factor; thus, the changes in mRNA steady-state levels may be an indirect consequence of the translational reprogramming.

**Figure 7.**
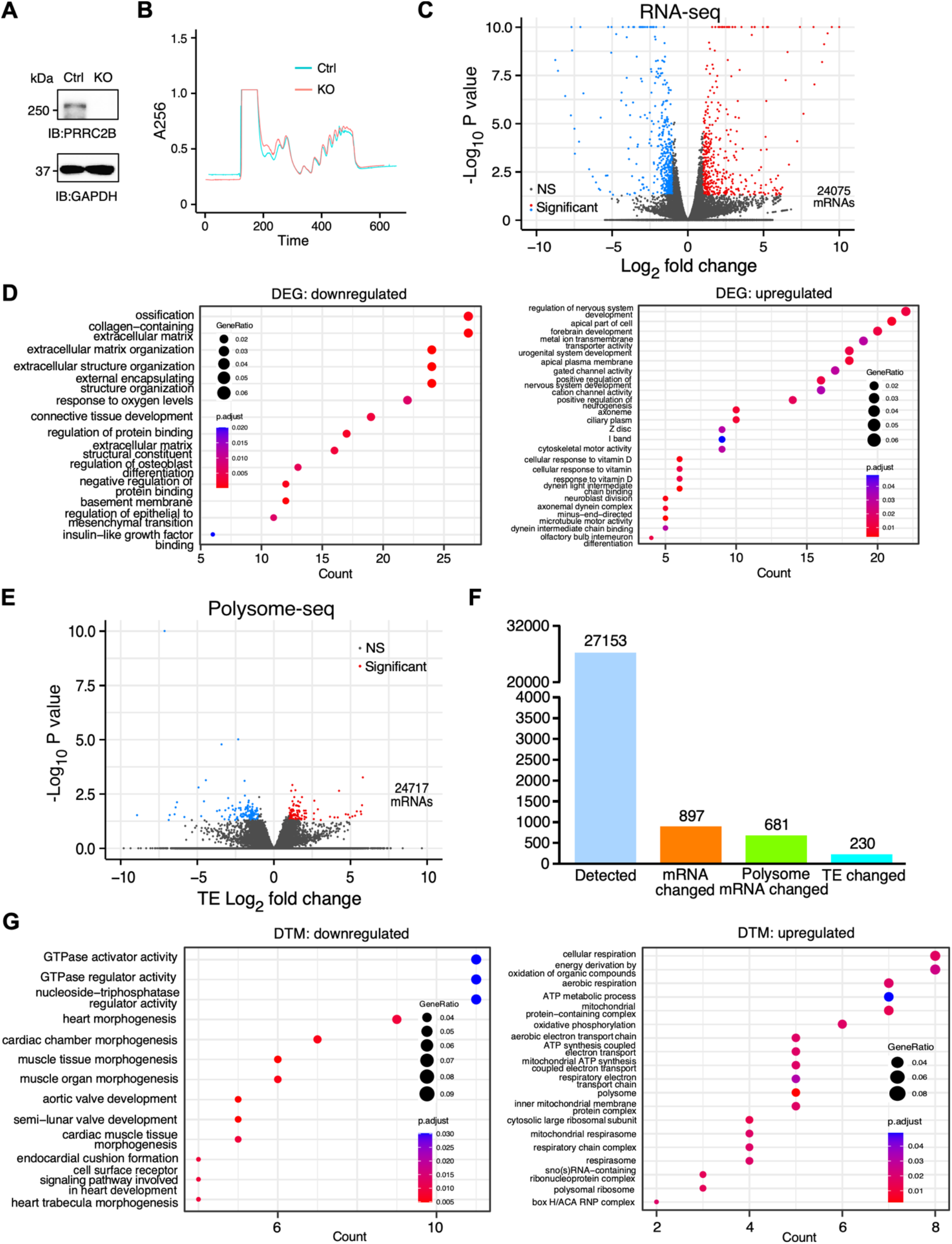
Transcriptomic and translatomic profiling of *PRRC2B* knockout human cells. **A**. Immunoblot validation of *PRRC2B* gene knockout in HEK293T cells. **B**. Polysome profiling of control and KO HEK293T cells. **C**. Volcano plot of differentially expressed genes identified by RNA-seq. **D**. GO analysis of significantly downregulated (left) and upregulated (right) genes. **E**. Volcano plot of differentially translated mRNAs (DTM) identified by polysome-seq. **F**. Number of genes significantly dysregulated at the RNA and translation efficiency (TE) levels in KO cells. **G**. Gene ontology analysis of significantly TE-downregulated (left) and -upregulated (right) mRNAs.

To further examine the translational reprogramming in *PRRC2B* KO human cells, we performed polysome profiling to isolate total RNA from the polysome fractions, followed by deep sequencing (polysome-seq). The polysome-seq analysis revealed PRRC2B-regulated target mRNAs when normalized to RNA-seq data to indicate the changes in translation efficiency (TE) based on the extent of enrichment of a given mRNA in actively translated polysome fractions (**Figure 7E, F, S7C-E; Table S7**). Intriguingly, TE-downregulated genes were highly enriched in cardiac development pathways, including aortic valve development, muscle tissue morphogenesis, semi- lunar valve development, cardiac chamber morphogenesis, endocardial cushion formation, heart morphogenesis, heart trabecula morphogenesis, and cardiac epithelial-to-mesenchymal transition, among others (**Figure 7G, left**). Multiple mRNAs enriched across various cardiac development pathways include *BMP2* (bone morphogenetic protein 2), *NOTCH1* (notch receptor 1), *RBPJ* (recombination signal binding protein for immunoglobulin kappa J region), *ROCK1* (Rho-associated coiled-coil containing protein kinase 1), *SNAI2* (snail family transcriptional repressor 2), *ANKRD1* (ankyrin repeat domain 1), *DSP* (desmoplakin), *MYLK* (myosin light chain kinase), *ADAMTS1* (ADAM Metallopeptidase With Thrombospondin Type 1 Motif 1), *DNAH11* (Dynein axonemal heavy chain 11), and *SOS1* (Son of sevenless homolog 1) (**Table S7**). Among these TE-downregulated proteins, the loss-of-function of RBPJ has previously been shown to cause PDA, whereas DA-enriched enzyme ROCK1 has been proposed as a therapeutic target for PDA (25). As a potential compensatory response, we observed that mitochondrial electron transport chain and ribosome-related mRNAs were enriched in the TE-upregulated cohort (**Figure 7G, right**).

We overlapped the proteins downregulated in the *Prrc2b*^tm1b-/-^ hearts with translational silenced genes in the KO HEK293T cells and inducible shRNA-mediated knockdown HEK293T cells we reported previously (9). We found 41 common hits among the three datasets, 134 shared genes between *PRRC2B* KO mouse hearts and KO human cells, and 16 additional shared genes between *PRRC2B* KO mouse hearts and knockdown human cells (**Figure 8A; Table S7**), suggesting a conserved cohort of PRRC2B-regulated mRNAs in different cell types across humans and mice. Among the 41 hits, more than half the genes encode metabolic enzymes, including nucleic acid metabolic enzymes CTP synthase 2 (CTPS2) and ribonucleotide reductase regulatory subunit M2 (RRM2), RNA-binding proteins leucine-rich pentatricopeptide repeat containing (LRPPRC) and Ro60, Y RNA binding protein (RO60), protein kinases such as ribosomal protein S6 kinase A5 (RPS6KA5) and mitogen-activated protein kinase kinase kinase kinase 5 (MAP4K5), NRAS proto-oncogene, GTPase (NRAS), ADP ribosylation factor guanine nucleotide exchange factor 2 (ARFGEF2), mitochondrial antiviral signaling protein (MAVS), phosphoribosyl pyrophosphate synthetase 2 (PRPS2), and lysine demethylase 3B (KDM3B). We then overlapped 191 genes downregulated in both *Prrc2b* mouse hearts (at the protein level) and translationally reduced in KO or knockdown human cells (at the TE level) with the previously reported PRRC2B-bound mRNAs, together with mouse PDA genes (**Figure 8B**). Seven mRNAs of *PLCG2* (phospholipase C gamma 2), *PHKB* (phosphorylase kinase regulatory subunit beta), *WDR26* (WD repeat domain 26), *XRCC6* (X-ray repair cross complementing 6; Ku70), *TMBIM6* (transmembrane BAX inhibitor motif containing 6; BI-1, BAX inhibitor 1), *PDE12* (phosphodiesterase 12), and *EMC3* (ER membrane protein complex subunit 3) are physical PRRC2B-interacting targets reduced at the protein level in *Prrc2b* KO hearts, as uncovered in this study.

**Figure 8.**
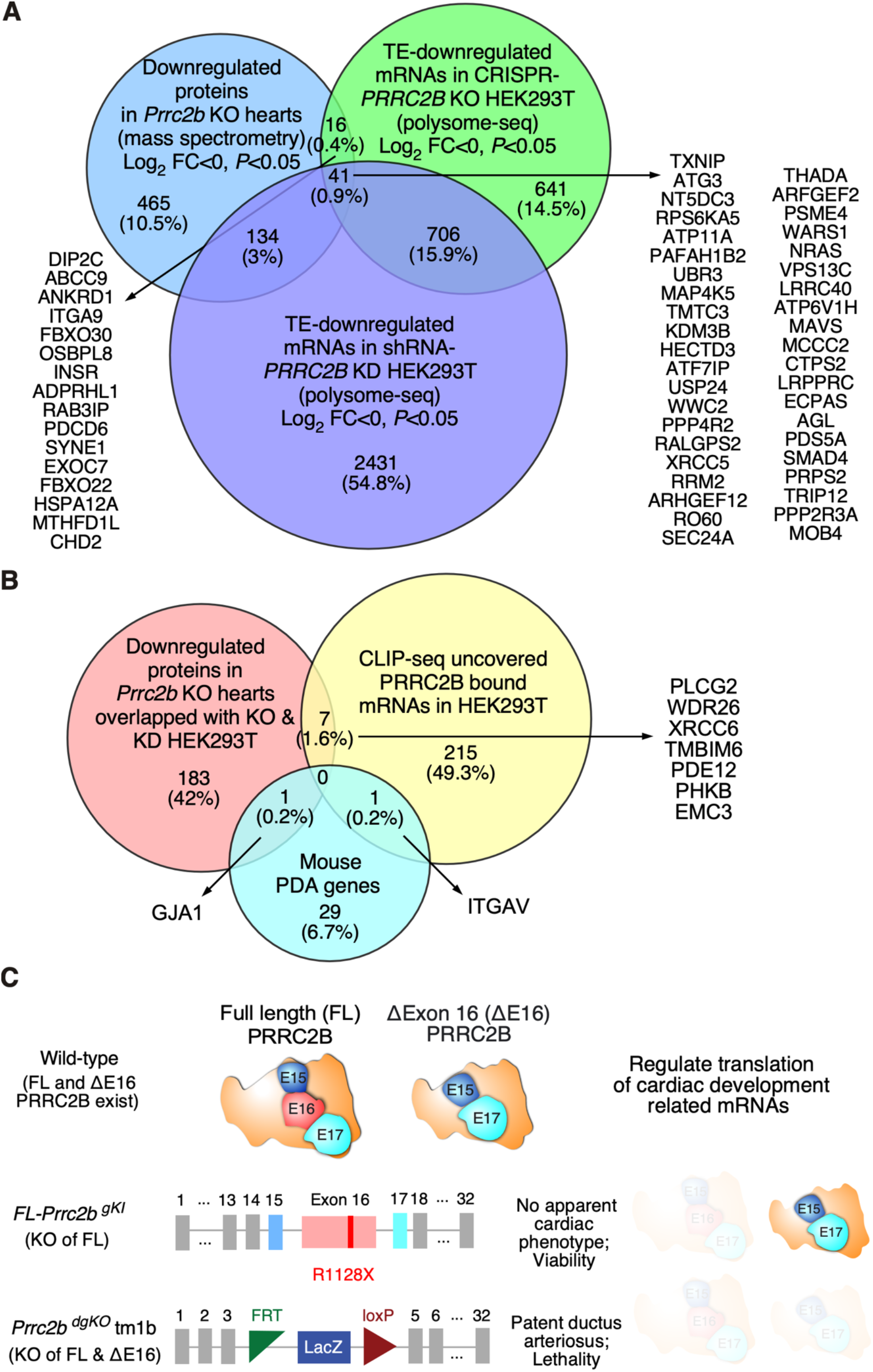
Conserved PRRC2B-regulated target mRNAs across cell types and species. **A**. Venn diagram of shared PRRC2B-regulated genes from *Prrc2b*^tm1b-/-^ gKO mouse hearts and *PRRC2B* KO and KD human HEK293T cells. Conserved downregulated genes at the protein translation level are listed. **B**. Venn diagram of overlapping reduced proteins in *Prrc2b*^tm1b-/-^ KO mouse hearts with PRRC2B-bound mRNAs from human cells and well-known mouse PDA genes. Conserved downregulated genes at the protein translation level are listed. **C**. Schematic model showing two alternative spliced *PRRC2B* isoforms (FL and ΔE16) and phenotypes of the *Prrc2b* genetic mouse models.

### Identification and comparison of the protein interactome of FL and ΔE16 PRRC2B isoforms and RNA-protein interacting domains

Using multiple genetic loss-of-function models in mice and human cells, we confirmed the essentiality of PRRC2B for basic cellular function and organismal development. However, the functional divergence between the full-length and ΔE16 alternative spliced isoforms remains poorly understood. To provide further unbiased datasets and mechanistic insights towards resolving this question, we first performed immunoprecipitation-mass spectrometry (IP-MS) analysis using a FLAG antibody after overexpressing the full-length and ΔE16 PRRC2B isoforms in HEK293T cells, with ribonuclease treatment (RNase A) to remove any RNA-mediated indirectly associated proteins (**Figure S8A, B, Table S8**). Both PRRC2B protein isoforms bind to the same set of translation initiation factors, including the noncanonical translational initiation factor eIF4G2, general essential translation initiation factor complex eIF3 components, among other translation machinery proteins, and translation-modulating RNA-binding proteins (such as ribosomal proteins, FXR1, among others) (**Figure S8A, B, Table S8**). Full-length PRRC2B has significantly more enriched protein partners localized in the nucleus, involved in DNA chromatin remodeling, gene transcription, and pre-mRNA splicing processes (**Figure S8C**). Consistently, the full-length PRRC2B protein was more localized in the nucleus than the ΔE16 isoform (**Figure S8D**).

Because *Prrc2b*^tm1b-/-^ KO mice, in which both FL and ΔE16 mRNA isoforms are depleted, exhibited PDA and early onset lethality due to aberrant cardiovascular development, whereas p.R1128X KI mice lacking only the FL mRNA isoform did not, we next explored the structure-function relationship of these two protein isoforms in their commonly shared domains and unique domains in the full-length protein. First, we performed RIP-RT-qPCR with a previously validated mRNA target *CCND2*, upon overexpression of the FLAG-tagged full-length PRRC2B, ΔE16, and N-terminal domains (1-450 aa or 1-600 aa) at comparable levels in HEK293T cells (data not shown). Full-length PRRC2B exhibited significantly higher binding ability with *CCND2* mRNA than the other three variants, suggesting the critical role of the exon 16-encoded ArgGly (RG)-rich RNA-binding region in interacting with PRRC2B target mRNAs in cells (**Figure S8E, F**). To further confirm the role of exon 16-encoded RG-rich RNA-binding motifs in interacting with PRRC2B target mRNAs, we introduced two independent mutant variants, Mut-1 and Mut-2, in which R^1065^GRGRG^1070^ and S^1106^GRGRG^1111^ were substituted with Ala (a stretch of AAAAAA), respectively (**Figure S8G**). RIP-RT-qPCR after overexpressing the FLAG-tagged full-length WT PRRC2B, and the two mutant protein variants showed a significant reduction in the protein-RNA interaction for three prior validated target mRNAs, *CRKL*, *CCND2*, and *YWHAZ* (**Figure S8H**). To prove the direct physical interaction between the exon 16-encoded domain and *CCND2* mRNA, we purified recombinant truncated PRRC2B protein (900-1200 aa containing the RG-rich RNA-binding regions) from bacterial strain BL21(DE3) (**Figure S8I**) and conducted electrophoresis mobility shift assay (EMSA) using *in vitro* transcribed *CCND2* mRNA 5’-UTR (100-nt). The EMSA analysis showed a dose-dependent formation of the protein-RNA complex (**Figure S8J**). These results confirmed the importance of exon 16 in maximizing the binding of the target mRNAs by PRRC2B, though the N-terminal domains are preserved for recruiting eIF4G2, eIF3, and the 40S ribosomal subunit to assemble the noncanonical translation initiation complex.

Although the FL PRRC2B contains the exon 16 that encodes the RG-rich RNA-binding regions, the genetic redundancy between the FL and ΔE16 isoforms (**Figures 2 and 3**) indicates that the shared ability in assembly of the eIF4G2-eIF3-containing translation initiation complex and partial compensation of mRNA-binding by other RNA-binding domains shared between the two isoforms. To confirm this essential activity of the N-terminal domains in recruiting the eIF4G2-eIF3 complex and determine the interaction sites in eIF4G2 for PRRC2B scaffolding, we overexpressed two major functional segments of the human eIF4G2 protein in HEK293T cells: 1-323 aa N-terminal and 545-890 aa C-terminal fragments. GST-tagged PRRC2B N-terminal fragment was expressed and purified from BL21(DE3) and could pull down the eIF4G2 C-terminal but not N-terminal fragment (**Figure S8K**). Overall, these results suggest that the ΔE16 alternatively spliced truncated isoform may partially compensate for the function of the full-length PRRC2B by preserving the capability of activating the eIF4G2-PRRC2B-eIF3 translation initiation complex pathway despite reduced binding affinity of the target mRNAs.

## Discussion

In this work, we identified two PRRC2B isoforms resulting from alternative splicing, including the full-length mRNA and a truncated isoform lacking exon 16 (ΔE16). Bioinformatic analysis of the PCGC database reveals multiple human PRRC2B mutations associated with CHD, including p.R1113X mutation, which is in exon 16. CRISPR-Cas9-mediated introduction of p.R1113X premature termination codon in the mouse genome does not cause any apparent cardiac abnormalities at the postnatal stage, possibly due to an intrinsic rescue by the alternative spliced isoform ΔE16. We further demonstrate that *Prrc2b* loss-of-function leads to PDA and, thus, perinatal lethality in mice (**Figure 8C**). Single-nucleus RNA-seq analysis of *Prrc2b* FL and ΔE16 double KO mice suggests changes in the number of SMCs and overall SMC gene expression in the whole heart, despite the absence of significant changes in gene expression in differentiated individual SMCs. Polysome-seq-based translatomic profiling in *Prrc2b* null hearts revealed *in vivo* PRRC2B-regulated target mRNAs encoding cell proliferation, migration, cellular metabolism, and SMC function-related proteins. Moreover, genetic knockout of the *PRRC2B* gene in human HEK293T cells reduces translation of mRNAs involved in heart and valve development, indicating a molecular-level translational regulatory function of PRRC2B, which is significantly associated with cardiovascular development.

Prior investigations of human cells and mouse models have indicated that alternative splicing is a primary intrinsic genetic rescue strategy employed by cells or tissues to counteract gene inactivation resulting from genetic manipulation (26). This finding is consistent with our observation that the human mutation mimicry R1128X KI mice exhibit no apparent cardiac phenotype, likely due to the rescue provided by ΔE16 isoform expression. Therefore, the R1113X human mutation is a modifier mutation (not a driver mutation) that contributes to congenital heart disease phenotypes. Alternative splicing of *PRRC2B* pre-mRNA generates two protein isoforms that may have potential functional redundancy that differentiates the phenotypes of *Prrc2b*^tm1b-/-^ versus *Prrc2b* p.R1128X KI mouse models. *Prrc2b* p.R1128X KI mice lost the FL PRRC2B isoform, while the ΔE16 isoform could compensate for the loss-of-function of the full-length protein, thereby maintaining vascular integrity and preventing PDA. This suggests that the ΔE16 isoform may partially or fully substitute the function of the FL protein *in vivo*. The evolution of this ΔE16 isoform may serve as a “fail-safe” strategy for the organism to maintain vascular well-being. Exon 16 encodes a domain with an RG repeat for RNA-binding, which may not be essential for organismal physiology in mice but is nevertheless required for optimized binding with target mRNAs in cells, based on our observations of viability in the *Prrc2b* p.R1128X KI mouse model.

This suggests that additional RNA-binding domains may still exist in PRRC2B. As a supporting hint, mRNA interactome capture analysis identified an mRNA-binding peptide from human PRRC2B (amino acids 385-398, located upstream of exon 16 spanning amino acids 775-1468), as mapped by mass spectrometry (7). This peptide is situated very close to the predicted alpha-helix regions (e.g., amino acids 350-370) by Alpha-Fold 2, while far away from the RG-rich domain in exon 16. As an alternative possibility, PRRC2B’s RNA-binding and complex-scaffolding functions may be compensated for by PRRC2A and PRRC2C, which possess the counterpart GR-rich RNA-binding domain and show high protein sequence similarity with PRRC2B. The functional divergence between the full-length and ΔE16 PRRC2B isoforms warrants future investigation, as indicated by others’ (19) and our proteomic interactome analyses.

Numerous studies have identified molecular clues required for highly coordinated cellular programs to mediate the complex regulation of DA (27). Notably, functional SMCs are vital for regulating postnatal DA closure after birth (28). To block the blood flow from the pulmonary artery to the aorta and facilitate full activation of pulmonary circulation, the DA vessel needs to be closed rapidly in a short time window after birth, requiring a temporary or spatial surge of protein synthesis for increasing SMC contractility or proliferation locally in the DA vessel areas but not in other blood vessels. When both ΔE16 and FL PRRC2B protein isoforms are absent in *Prrc2b* homozygous KO (tm1b) hearts, SMCs at the DA region may be affected due to compromised translation of specific mRNAs, leading to PDA. The proteomic changes in *Prrc2b* KO hearts suggest translational defects in a selective cohort of proteins triggered by PRRC2B loss of function (**Figure 6, 8A, B**). This finding is consistent with the notion that PRRC2B regulates the translation of a specific group of mRNAs in human cells, as previously reported by us (9). Several large-scale unbiased analyses have reported that PRRC2B is an RNA-binding protein that can interact directly with mRNA (5–7). Moreover, previous work from Dr. Yamanaka’s group and our laboratory has uncovered a protein complex of PRRC2B with translation initiation factor eIF4G2 and the eIF3 complex in mouse embryonic stem cells and HEK293T cells, as determined by mass spectrometry (8,9), suggesting a conserved composition of PRRC2B-containing complexes involved in transcript-selective translational control across different cell types. Our current work on *Prrc2b* KO mice indicates that PRRC2B may play an essential role in cardiac vessel development by regulating temporally and spatially restricted mRNA translation required for optimal proliferation or contraction in ductal SMCs and possibly other cardiac cell types. Our polysome-seq-based translatomic profiling of E18.5 *Prrc2b* KO and WT control hearts revealed convergent downstream pathways regulated by PRRC2B at the translational level, including cell proliferation, migration, and cardiovascular function-related genes, compared with PRRC2B KO human cells. Consistent with this idea, *PRRC2B* null HEK293T cells show reduced translation of cardiac development-related mRNAs (*BMP2*, *NOTCH1*, *RBPJ*, *ROCK1*, *SNAI2*) (**Figure 7**), highlighting a conserved regulatory program across humans, mice, and different cell types. These findings support a mechanistic link between aberrant cardiovascular development and dysregulated mRNA translation.

Our transcriptomic profiling analysis of E18.5 prenatal *Prrc2b* KO hearts revealed downregulation of *Myh11* and *Smarca4* following *Prrc2b* deletion. Since both genes were essential for functional SMCs to orchestrate DA closure, SMC-specific *Smarca4* KO and *Myh11* global KO mice showed a prevalence of PDA due to insufficient SMC differentiation (*Smarca4* KO) or reduced SMC contractility (*Myh11* KO), respectively (29,30). Also, diminished expression of DA closure-related genes (*Acta2*, *Actg2*) responsible for the vascular tone of SMCs (31) was evident in the heart of *Prrc2b*^tm1b-/-^ mice. However, the gene dysregulation at the mRNA level is likely either a secondary effect downstream of mRNA translational changes or due to another function of PRRC2B in regulating mRNA stability (11). Moreover, snRNA-seq data indicate reduced SMC proliferation or cardiac progenitor cell-to-SMC differentiation. Therefore, we infer that reduced prenatal overall expression of DA-regulating genes upon *Prrc2b* deletion and decreased number of SMCs in the heart may also contribute to the PDA phenotype after birth, in addition to compromised mRNA translation.

Taken together, we found that the loss of function of two alternative splicing isoforms of PRRC2B simultaneously caused CHD (**Figure 8C**). PRRC2B is a novel translation regulatory factor related to cardiovascular development and CHD. Evolving two alternatively spliced isoforms of PRRC2B may be essential for maintaining cardiovascular integrity and species survival. This work provides novel insights into the interplay among PRRC2B alternative splicing, PRRC2B-mediated translational control, and congenital cardiovascular disorders. Our findings may shed light on the significance of PRRC2B in future diagnosis and treatment of human cardiovascular disease.

## Materials and methods

### Whole exome sequencing and burden testing for published PCGC databases

Results of genomic analysis using the Pediatric Cardiac Genomic Consortium (PCGC) whole-exome cohort have been previously published (21); the study presented here follows previously described protocols (13,32,33). Briefly, whole-exome DNA from blood or salivary samples was captured using the Nimblegen v.2 exome capture reagent (Roche) or Nimblegen SeqxCap EZ MedExome Target Enrichment Kit (Roche), followed by Illumina DNA sequencing as previously described (13,32,33). The generated data were processed at the Yale University School of Medicine, and reads were mapped to the hg19 reference genome. Mapped reads were further processed using the GATK Best Practices workflows (34), as previously described (32). Single-nucleotide variants and small indels were called with GATK HaplotypeCaller (35). Further filtering of the data set was performed using PLINK (36), including the removal of individuals with low call rates, outlying heterozygosity rates, outlying relatedness rates, and sex discrepancies. A total of 3,740 probands passed individual filtering (21). Variants were filtered based on call rate, Hardy-Weinberg equilibrium, and a high number of Mendel errors (≥3). Variants within PRRC2B were extracted using the longest known canonical hg19 PRRC2B variant BED file downloaded from UCSC Table Browser on 9/19/2021 (37). The remaining variants were annotated using ANNOVAR (38). Loss-of-function (frameshift, splice-site, start-lost, stop-gained, and stop-loss) and missense variants with a CADD (39) score >20 and a minor allele frequency <0.001 across all gnomAD (40) populations were included in the burden analysis. Dichotomous traits were created by comparing probands with a specific phenotype to the rest of the probands in the PCGC whole-exome cohort. Burden testing for these dichotomous traits was performed using the sequence kernel association test (SKAT) (41) package in R, with the first three principal components as covariates.

### Mice

All mouse experiments were conducted in accordance with protocols approved by the University Committee on Animal Resources (UCAR) of the University of Rochester Medical Center (URMC). The mice were housed in a 12:12 hours light: dark cycle in a temperature–controlled room in the animal housing room at URMC, with free access to water and food. Both male and female mice aged 8-12 weeks were used. All the mice are on a C57BL/6J background. Carbon dioxide euthanasia is used for terminating mice. All anesthetic and analgesic agents used in the study were approved by the UCAR of the URMC.

### Generation of *Prrc2b* knockout mice (*Prrc2b*^tm1b-/-^)

*Prrc2b*^tm1b-/-^ mice were generated using the knockout-first strategy. A detailed schematic representation of the targeted *Prrc2b* allele is illustrated in Figure 3A. The *Prrc2b*-targeting vector (PG00197_Z_2_F04) was obtained from the Knockout Mouse Project Repository (IKMC project: 71689). Heterozygous knockout-first (*Prrc2b*^tm1a^) mice containing a promoter-driven cassette (L1L2_Bact_P), which was inserted at position 32074888 of Chromosome 2 upstream of the critical exon four of the *Prrc2b* gene. Then, *Prrc2b^flx^* tm1a mice were mated with Tg (CMV-Cre) mice to generate an exon four knockout allele (*Prrc2b*^gKO^ tm1b) (**Figure 3**). The PCR primers used for genotyping to validate the knockout alleles are summarized in Table S9.

### Generation of *Prrc2b* R1128X knock-in mice

*Prrc2b* global R1128X knock-in (KI) mice were generated by the Mouse Genome Editing Resource at URMC using the CRISPR-Cas9 method. The guide RNA (gRNA) used for *Prrc2b* KI was designed using an online CRISPR RNA design tool developed by Dr. Feng Zhang’s lab (http://crispr.mit.edu). The gRNA with the highest score and lowest off-target probability was chosen. The efficiency of gRNA and Cas9-2xNLS (Synthego) was tested *in vitro* in a tube using a 500-bp mouse genomic DNA-derived PCR product containing the target sequence. Then, 25 pmol of single guide RNA (sgRNA) and 25 pmol of Cas9 nuclease were mixed in the injection buffer in a 12.5 μL reaction. The mixture was then incubated at room temperature for 10 minutes to form the RNP complex. Finally, this RNP complex was mixed with a single-strand DNA template that contains an Arg codon-to-stop codon at the 1128^th^ amino acid position and two additional wobble base mutations (GACTTC-to-GA<u>T</u>TT<u>T</u>) at a 1:3 molar ratio for pronuclear injection. In this project, injected embryos were transferred into multiple recipient C57BL/6J mice, yielding mosaic pups carrying the desired mutations. To obtain heterozygous gKI mice for experiments, injected mice were bred with C57BL/6J WT mice to achieve germline transmission. They bred with WT mice for 5 generations to eliminate potential off-target sites. Genotyping was performed by Sanger sequencing of PCR-amplified fragments spanning the WT versus the gKI mutation regions.

Guide (g)RNA (20 nt):

5’- CCCUGCCUCGGUUACUGCGC -3’

ssDNA template (194 nt; 5’ – 3’):

GCAGGCAGCAGCACAAGTGGTCTTTGTGGCACGGGTGTCCTTGGGTCTCGTGGCATGTAC

AGTAGTGGGCAGCGCAGTAACCGAGGCAGGGGCCTG<u>TGAGATTTT</u>CCCCCACCAGAAGA

CTGCCCCAGAGCCAAGCCAAGGCGTCGCATTGCCAGCGAGACTCACAGCGAGGGCTCTGAGTATGAAGAGCTGCC

### Perinatal mouse heart isolation

*Prrc2b*^tm1b+/-^ mice were set up for timed mating in the evening to harvest embryos from the exact day of gestation, ranging from E12.5 (a prenatal stage with a developed heart) to P0 (an early postnatal stage). Vaginal plugs (white to yellowish ejaculate) were checked early in the morning to confirm pregnancy. Once pregnancy was confirmed, their body weights were recorded regularly until the desired harvest stage was reached. For prenatal stage harvesting, the pregnant mice were euthanized with a mixture of ketamine and xylazine via intraperitoneal injection. Individual embryos were extracted from the uterus, followed by heart isolation under the dissection microscope. They were immersed in prechilled 1x phosphate-buffered saline (PBS) to wash out excess blood before isolating the hearts. For harvesting at P0, pregnant mice were closely monitored to determine the exact delivery date and time. Then, the freshly delivered fetuses were separated and euthanized with rapid freezing. The mouse sternum was dissected, and the heart was exposed before being perfused with 1x PBS buffer and then isolated.

### Echocardiography

Echocardiographic image collection was performed using a Vevo2100 echocardiography machine (VisualSonics, Toronto, Canada) and a linear-array 40 MHz transducer (MS-550D) by the surgical core of Aab Cardiovascular Research Institute at URMC in a blinded manner. Image capture was performed in mice under general isoflurane anesthesia with heart rate maintained at 500-550 beats/min. LV systolic and diastolic measurements were captured in M-mode from the parasternal short axis. Echocardiographic images were collected at baseline and 28 days after Ang II and PE infusion, and monthly in untreated mice during natural aging, from both sexes, and analyzed. Hearts were harvested at multiple endpoints in different experiments.

### Cell culture and transfections

Human HEK293T cells (ATCC, Cat. #CRL-3216) were propagated in Dulbecco’s modified Eagle’s medium (DMEM; Corning) supplemented with 10% fetal bovine serum. Cells were transfected with plasmids using Lipofectamine 3000 (Invitrogen) according to the manufacturer’s instructions. Human aorta-derived smooth muscle cells (HASMCs) (Cell Applications, Cat. #354-05a) with passage number lower than 8 (P8) were cultured and passaged in All-in-one ready-to-use Human Smooth Muscle Cell Media (Cell Applications, Cat #311-500).

### Molecular cloning

DNA fragments were obtained and amplified by PCR from genomic DNA or cDNA extracted from HEK293T cells using TRIzol (ThermoFisher) according to the manufacturer’s instructions. Amplified DNA fragments were separated on a 1% agarose gel, purified, digested by restriction endonucleases together with the backbone plasmid at 37°C overnight, and ligated to the cleaved backbone by T4 DNA ligase (NEB). The ligated products were transformed into *E. coli* competent cells (DH5α). Miniprep was performed on single colonies to purify the plasmids. Plasmid sequences were then confirmed by Sanger sequencing in Azenta.

### Total RNA extraction

Total RNAs were harvested from the heart tissues of neonatal mice (P0) using TRIzol reagent (ThermoFisher). Briefly, the heart tissues were homogenized in 500 μL of TRIzol for 1 minute using the Precellys kit (Bertin Technologies) on the Minilys Personal Homogenizer (Bertin Technologies). Then, the homogenates were placed on ice for 10 minutes to allow for complete lysis. Then, 100 μL of chloroform was added to the homogenates, followed by vigorous hand- shaking for 15 seconds. The mixture was then incubated at room temperature for 5 minutes. The mixture was centrifuged to separate the lower phenol: chloroform phase, an interface, and an upper aqueous phase. The upper layer was transferred to a fresh tube, followed by the addition of 500 μL of isopropanol and 2 μL of 20 mg/mL glycogen for precipitation. After centrifugation, the pellet was washed once with 75% ethanol, then centrifuged again to obtain the RNA pellet.

### mRNA expression by agarose gel electrophoresis of cDNA after PCR or qPCR

After RNA isolation, 1 μg of total RNA was used as the template for reverse transcription using SuperScript^TM^ IV RT (Invitrogen) following the manufacturer’s instructions. cDNA was then generated to detect *Prrc2b* mRNA expression at different exons, followed by PCR. *Gapdh* was used as a housekeeping control. Then, the PCR products were loaded in a 1.5% agarose gel and run at 140 V for 30 minutes before visualization. For qPCR, the procedure is as follows: 1) initial denaturation at 95°C for 60 seconds. 2) 40 cycles of denaturation at 95°C for 10 seconds and annealing/extension at 60°C for 45 seconds. 3) melt curve analysis by 0.5°C increments at 5 seconds / step between 65-95°C.

### Flow cytometry

The embryonic hearts were harvested as described above. Briefly, the pregnant mice were euthanized with a mixture of ketamine and xylazine via intraperitoneal injection. Individual embryos were extracted from the uterus, followed by heart isolation under the dissection microscope. They were immersed in 1x phosphate-buffered saline (PBS) to wash out excess blood before isolating the hearts. Individual hearts were then digested with collagenase (0.5 mg/ml) supplemented with 1x PBS at 37°C for 10 minutes using a magnetic stirrer. Upon complete digestion, the cell suspension was neutralized in PBS containing 2% fetal bovine serum (FBS), then washed with 1x PBS. For analysis of cell-surface markers, cells were first incubated with TruStain FcX™ (anti-mouse CD16/32) antibody (BioLegend, Cat. #101319) and viability dye (LIVE/DEAD™ Fixable Aqua Dead Cell Stain Kit, Cat. #L34966) diluted in PBS for 30 minutes at 4°C in the dark to block non-specific binding of immunoglobulin to mouse Fc receptor and differentiate live and dead cells, respectively. Following staining, cells were washed twice with staining buffer. Then fixed using the Ebioscience™ Foxp3/Transcription Factor Staining Kit (ThermoFisher, Cat. #00-5523-00) according to the manufacturer’s protocol. Cells were incubated with fluorochrome-conjugated antibodies (listed in the supplemental file) for 30 minutes at 4°C in the dark for intracellular staining. Perm/Wash buffer used to wash the cells, followed by resuspension in staining buffer (ThermoFisher, Cat. #00-4222-26) for acquisition. Data were acquired on a BD Biosciences LSRFortessa, and at least 10,000 live events were collected per sample. Individual antibody-specific isotype controls and unstained controls were used for appropriate gating. Data were analyzed using FlowJo software. Cell populations were identified by sequential gating to exclude debris and doublets, then gating on Live cells and on individual marker-positive populations.

### Cell proliferation and migration quantification

According to the manufacturer’s instructions, the cell growth and proliferation rates of human aorta-derived smooth muscle cells (HASMCs) with or without lentiviral-shRNA-mediated knockdown of PRRC2B were determined in a 96-well plate using the Cell Proliferation Kit I (MTT-based) colorimetric assay (Roche, Cat. #11465007001). Meanwhile, HASMC migration upon PRRC2B knockdown compared to the control scrambled shRNA treatment was evaluated using Biocoat cell culture inserts in a transwell migration assay without collagen 1 coating. FBS were used as a chemoattractant to evaluate the cell migration rate.

### Immunofluorescence (IF) and Wheat Germ Agglutinin (WGA) staining

For heart tissue sections, paraffin-embedded human heart tissue or mouse whole-heart sections were deparaffinized and hydrated using an automated deparaffinization and hydration system. After unloading from the machine, the slides were incubated in water, then antigen-retrieved in 10 mM sodium citrate buffer. Following a 5-minute wash with 1x PBS, the slides were quenched with a 3% hydrogen peroxide (H_2_O_2_) solution in 1x PBS for 30 minutes. The slides were then permeabilized with 0.1% Triton X-100 in PBS for 8 minutes, blocked with 5% goat serum in PBS for 1 hour at room temperature, and incubated overnight with primary antibodies. After washing off the primary antibody with 1x PBS, appropriate secondary antibodies (anti-rabbit IgG-Alexa Fluor 594, Cat. #A11012, 1:200; anti-mouse IgG-Alexa Fluor 488, Cat. #A11029, 1:200; anti-mouse IgG-Alexa Fluor 594, Cat. #A11005, 1:200; anti-rabbit IgG-Alexa Fluor 488, Cat. #A11034, 1:200) was added and incubated for 45 minutes at room temperature. Finally, the slides were stained with DAPI-containing antifade mounting media (VECTASHIELD, Cat. #H-1500) before being covered with a coverslip. Images were acquired using an IX81 confocal microscope (Olympus).

For cells grown on coverglass or chambered slides, HEK293T cells were grown on the coverslips and transfected with a FLAG-tagged PRRC2B expression construct for 24 hours at 37°C before fixation for 10 minutes with 4% paraformaldehyde in 1x PBS. Cells were washed 3 times with 1x PBS for 5 minutes each, then permeabilized with ice-cold 0.5% Triton X-100 in PBS for 5 minutes. After blocking with 1% BSA in PBS, the coverslips were incubated with the indicated primary antibody (anti-FLAG, 1:2000) in blocking solution (2% BSA in PBS) for 1 hour at room temperature, then washed 3 times for 5 minutes each with PBS. The coverslips were incubated with Alexa Fluor-488-conjugated secondary antibodies (ThermoFisher, 1:1000) in PBS and washed 3 times for 5 minutes each in PBS. Coverslips were air-dried and placed on slides with antifade mounting medium (containing DAPI). The slides were imaged using an Olympus FV1000 confocal microscope. A negative control using only the same amount of secondary antibody was performed for IF or IHC to confirm the fluorescence signals as genuine target staining and not non-specific background.

The following steps were performed to stain the heart tissue section with WGA. First, a 3% H_2_O_2-_ quenched heart tissue section was incubated with 10 μg/ml of WGA-AF488 (Invitrogen, Cat. #W11261) in 1x PBS for 1.5 hours at room temperature in the dark. Next, the slides were rinsed 3 times with 1x PBS-T (0.5% Tween 20) for 5 minutes each, followed by a final wash with 1x PBS. Finally, the slides were stained with DAPI-containing antifade mounting media (VECTASHIELD, Cat. #H-1500), and a coverslip was applied. Images were acquired using an upright Epifluorescent microscope (BX51, Olympus).

### Picrosirius red staining

The heart tissue sections were stained with Picrosirius Red Solution (Abcam, Cat. #ab246832) with minor modifications. An adequate amount of the solution was applied to cover the tissue section, and the section was incubated for 60 minutes. The sections were then quickly rinsed twice with 0.5% acetic acid solution and once with absolute alcohol. Finally, a coverslip was mounted on the slides using Cytoseal^TM^ XYL mountant (Epredia^TM^, Cat. #8312-4) before microscopic examination.

### Co-immunoprecipitation

Cells from one 80% confluent 10-cm culture dish were harvested using a native lysis buffer (300 μL) with 50 mM HEPES, pH 7.5, 150 mM KCl, 2 mM EDTA, 1 mM NaF, 0.1 % (v/v) Nonidet P-40, 1 mM DTT, and 25 μl/ ml protease inhibitor cocktail (Sigma-Aldrich) for mammalian tissues. RNasin RNase inhibitor (Promega) or RNase A was added for RNA-dependent and independent interactome capture. Immunoprecipitation (IP) of endogenous PRRC2B was performed using 1 μg (dilution factor 1:125) rabbit anti-human PRRC2B antibody (Invitrogen), followed by Magnetic dynabead protein A (Invitrogen) pull-down. IPs of FLAG-tagged PRRC2B and PRRC2B fragments were performed using anti-DYKDDDDK Magnetic agarose (ThermoFisher). Beads were mixed with 70 μl of SDS loading buffer (Bio-Rad) and boiled for 5 min to elute proteins for Western blot and mass spectrometry analysis.

### Mass spectrometry analysis

Mass spectrometry (MS) was performed by the Mass Spectrometry Resource Lab of the University of Rochester Medical Center. Briefly, the IP elution was loaded onto a 4-12% gradient SDS-PAGE gel, run for 10 minutes, and stained with SimplyBlue SafeStain (Invitrogen). The stained regions were then excised, cut into 1 mm cubes, de-stained, reduced, and alkylated with DTT and IAA (Sigma), dehydrated with acetonitrile, and incubated with trypsin (Promega) at 37°C overnight. Peptides were then extracted by 0.1% TFA and 50% acetonitrile, dried down in a CentriVap concentrator (Labconco), and injected onto a homemade 30 cm C18 column with 1.8 μm beads (Sepax) on an Easy nLC-1000 HPLC (ThermoFisher) connected to a Q Exactive Plus mass spectrometer (ThermoFisher) for MS. Raw data from MS experiments were mapped against the SwissProt human database using the SEQUEST search engine within the Proteome Discoverer software platform, version 2.2 (ThermoFisher). The Minora node was used to determine relative protein abundance using the default settings. Percolator was used as the FDR calculator, and peptides with q-values> 0.1 were filtered out. Biological triplicates and three IPs using rabbit pre-immune IgG (negative control) were included for both quantitative proteomic analyses comparing E18.5 WT and *Prrc2b* gKO mouse hearts and IP-MS in HEK293T cells after overexpression of FL and spliced isoforms of PRRC2B.

### Western blotting

Cells were lysed in RIPA buffer (ThermoFisher), and total cellular proteins were separated in a 10% denaturing polyacrylamide gel, transferred to polyvinylidene difluoride membranes (PVDF; Amersham Biosciences), probed using primary antibodies, and incubated with either a mouse or rabbit secondary antibody conjugated with horseradish peroxidase (GE Biosciences). According to the manufacturer’s suggestions, protein abundance was quantified by the Pierce BCA Protein Assay kit (ThermoFisher, Cat. #UL294765) for quantitative Western blotting. If applicable, the same protein mass was loaded, and β-actin was used as an internal control.

### RNA-binding protein immunoprecipitation (RIP)

Dynabeads-Protein A magnetic particles (Invitrogen, Cat. #3201250) were incubated with rabbit anti-FLAG IgG (Biotec, Cat. #20543-1-AP) or rabbit normal IgG (Cell Signaling, Cat. #2729S) for 2 hours at 4°C. For one pellet, 12 μL of the beads and 2.2 μg of the antibodies were used. Unbound IgG was washed out with NET2 Buffer with 50 mM Tris-HCl, pH 7.5, 150 mM NaCl, and 0.5% NP-40. Cells from one 80% confluent 10-cm culture dish were washed and harvested in ice-cold PBS, followed by a spin down at 500x g for 5 minutes at 4°C. Cell pellet was incubated for 10 minutes on ice in hypotonic lysis buffer (220 μL) with 10 mM Tris-HCl (pH 7.5), 10 mM NaCl, 10 mM EDTA, 0.5 % (v/v) Triton X-100, 0.03 units/μL RNase inhibitor (NEB), and 1x Protease Inhibitor (Roche). Then 18 μL of 4 M NaCl was added. The lysate was clarified by spin down at 21,000x g for 10 minutes at 4°C. 12 μL of the supernatant was taken as pre-IP input fraction and treated with Trizol. The rest of the lysate was pre-cleaned with 22 μL of Dynabeads-Protein A magnetic particles (Invitrogen, Cat. #3201250) in HLB for 2 hours at 4°C and clarified by spin down at 15,000x g for 10 minutes at 4°C. 30 μL of the lysate was taken for WB. The rest of the lysate was split by 100 μL and incubated with 12 μL of the reassociated Dynabeads-anti-FLAG IgG or Dynabeads-Normal IgG for 2 hours at 4°C. Supernatant was discarded, and the beads were washed 3 times with NET2 buffer. The beads were split into two equal parts for elutions. Protein elution was performed by incubating the beads in 30 μL Laemmli sample buffer for 10 minutes at 95°C. RNA elution was performed in 25 μL of HLB with 1% SDS and 1.2 μg/μL Proteinase K (NEB) for 30 minutes at 55°C. RNA extraction and reverse transcription were performed as described above with minor modifications. 250 ng of total RNA was used as a template for reverse transcription. qPCR of 2 μL of cDNA was performed in iTaq™ Universal SYBR® Green Supermix (BioRad, Cat. #1725124) premix with primers for CCND2 (hCCND2-f, hCCND2-r), YWHAZ (hYWHAZ-f, hYWHAZ-r), ACTB (hACTB-f, hACTB-r) transcripts, and 18S RNA cDNA (18srRNA-f, 18srRNA-r) as a control. The Ct (cycle threshold) of each cDNA was normalized to the 18S signal, and enrichment was calculated as the ratio of anti-FLAG IP samples to normal IgG IP samples.

### Protein expression and purification of recombinant proteins of PRRC2B and eIF4G2 fragments using nickel-based immobilized metal affinity chromatography

Fragments of eIF4G2 1-323, 48-323, 540-897 (aa) were PCR amplified from pcDNA3-HA-eIF4G2 (whole plasmid sequence verified by Plasmidosauris), cloned with NdeI and XhoI sites in modified pET28a vector with His-tag and TEV cleavage site downstream. A fragment of PRRC2B 1-600 (aa) was PCR-amplified from pcDNA3-FLAG-PRRC2B and cloned into the BamHI and EcoRI sites of the pGEX-4T1 vector. One liter of BL21(DE3) (ThermoFisher, Cat. #EC0114) *E. coli* bacteria culture was used for overexpression. Protein expression was induced with 0.4 mM IPTG (GoldBio, Cat. #2481C) at an optical density (OD) of 0.6, and the bacteria were incubated on a shaker at 20°C for 12 hours. The cells were collected by centrifugation at 3000g for 15 minutes at 4°C, then resuspended in ice-cold lysis buffer 1 (20 mM HEPES-KOH, pH 7.5, 200 mM KCl, 20 mM imidazole, 0.05% Triton X-100, 0.7 mM PMSF, 5% glycerol) for eIF4G2 fragments or lysis buffer 2 (20 mM HEPES-KOH, pH 7.0, 300 mM KCl, 0.05% Triton X-100, 0.7 mM PMSF, 0.1 mM EDTA, 5% glycerol, 1x Protease inhibitor (Roche)) for GST-PRRC2B 1-600 or GST and lysed by French-press (1100 psi). The lysates were centrifuged at 14,000g for 30 minutes, and the supernatant was loaded onto Ni-NTA resin. The resin was washed with 80 mL of washing buffer 1 (20 mM HEPES-KOH, pH 7.5, 300 mM KCl, 40 mM imidazole, 0.7 mM PMSF, 5% glycerol) for eIF4G2 fragments or washing buffer 2 (20 mM HEPES-KOH, pH 7.0, 300 mM KCl, 0.7 mM PMSF, 0.1 mM EDTA, 5% glycerol) for GST-PRRC2B 1-600 or GST. The eIF4G2 fragments were eluted with the elution buffer 1 (20 mM HEPES-KOH, pH 7.5, 200 mM KCl, 300 mM imidazole, 5% glycerol, 1 mM DTT). GST-PRRC2B 1-600 and GST were eluted in elution buffer 2 (20 mM HEPES-KOH, pH 7.5, 200 mM KCl, 0.1 mM EDTA, 5% glycerol, 20 mM Glutathione (Thermo Scientific Chemicals, Cat. #AAA1801406), then concentrated. During concentration, the elution buffer was exchanged for the storage buffer (20 mM HEPES-KOH, pH 7.0, 100 mM KCl, 5% glycerol, 1 mM DTT; imidazole or glutathione removed from the buffer [<1 mM/0.05 mM]). The protein samples were resolved by PAGE, followed by Coomassie G-250 staining (VWR, Cat. #0615) to visualize the purified protein.

### GST-PRRC2B 1-600 and His6-eIF4G2 fragments pulldown assay to detect interaction

Fragments of eIF4G2 1-322, eIF4G2 545-890 were incubated with GST-PRRC2B 1-600 or GST as a control in binding buffer with 20 mM HEPES-KOH pH 7.0, 100 mM KCl, 0.025% Tween 20, 1 mM DTT, 5% glycerol, 1x Protease inhibitor for 15 minutes at 37°C. The GST-tag was captured using glutathione magnetic beads for 10 minutes at 4°C. The beads were washed with 50 μL of the binding buffer 3 times. Elution was performed in 20 mM HEPES-KOH, pH 7.0, 100 mM KCl, 0.025% Tween 20, 10 mM glutathione, pH 8.0, 5% glycerol for 10 minutes at 4°C. Fractions were analyzed by Western blotting with mouse anti-His (Proteintech; 1:1000) antibody.

### RNA electrophoretic mobility shift assay (EMSA)

Interactions between PRRC2B protein fragments and *CCND2* 5’-UTR RNA (*in vitro* transcribed by T7 RNA polymerase) were evaluated by RNA EMSA. Pre-run was performed for 30 minutes (100 V) for native PAGE gel containing 8% acrylamide, 10% glycerol, 1x TBE (pH 8.1-8.3). 2 μM RNA was annealed at 66°C for 2 minutes in binding buffer (20 mM HEPES-KOH pH 7.5, 100 mM KCl, 1.5 mM MgCl_2_, 0.01% Tween 20, 1 mM DTT, 5% glycerol) then cool on bench. Protein at different concentrations (1, 2, and 5 μM) and RNase inhibitor (1 U) were added, and the mixture of 20 μL of reaction was incubated for 10 minutes at 37°C. Subsequently, the loading buffer (bromophenol blue, 10% glycerol, and binding buffer) was added, and the sample was loaded into the gel. The RNA–protein complex samples were resolved by 8% native PAGE for 2.5 hours at 100 V. The gel was stained with 1:10,000 SYBR Gold in TBE for 10 minutes, then washed in TBE for 20 minutes.

### Polysome profiling and RNA extraction

E18.5 mouse embryonic hearts were snap frozen in liquid nitrogen, or 1 × 10^9^ HEK293T cells were incubated with cycloheximide (100 μg/ml; Sigma) for 10 minutes. The heart tissues or cells were then harvested using a native lysis buffer with 100 mM KCl, 5 mM MgCl_2_, 10 mM HEPES, pH 7.0, 0.5% Nonidet P-40, 1 mM DTT, 100 U/ml RNasin RNase inhibitor (Promega), 2 mM vanadyl ribonucleoside complexes solution (Sigma), 25 μl/ ml protease inhibitor cocktail (Sigma), cycloheximide (100 μg/ml). The lysate was spun down at 1,500g for 5 minutes to pellet the nuclei. The supernatant was loaded onto a 10-50% sucrose gradient and spun in an ultracentrifuge at 150,000g for 2 hours. The gradients were then transferred to a fractionator coupled to an ultraviolet absorbance detector that outputs an electronic trace across the gradient. Using a 60% sucrose chase solution, the gradient was pumped into the fractionator and divided equally into 9 or 10 fractions. For subsequent RNA extraction, 400 μL of each fraction was mixed with an equal volume of chloroform: phenol: isoamyl alcohol and 0.1x 3M sodium acetate (pH 5.2), then spun down at 16,000g for 10 minutes. The upper aqueous layer was mixed with 3 volumes of 100% ethanol and 0.1 volume of 3 M sodium acetate (pH 5.2), then incubated at -20°C overnight. The solution was then spun at maximum speed for 30 minutes to pellet the RNA, which was washed twice with 70% ethanol and resuspended in nuclease-free water. DNase I was then added to remove genomic DNA contamination, followed by another round of RNA purification. For RNA deep sequencing, RNA purified from fractions #5-9 was combined into a single polysome-associated RNA sample, which was then sequenced (polysome-seq) or followed by RT-qPCR. DESeq2 was used to test the RNA fold changes and their significance between KO and WT heart samples. For polysome-seq, translation efficiency (TE) fold changes were calculated from normalized gene expression counts for E18.5 WT control hearts and *Prrc2b* global KO hearts, respectively. The Wilcoxon test was used to compare the WT and KO groups and assess significance. The significance of the TE fold-change is based on the *p*-value.

### RNA-sequencing analysis and data processing

As aforementioned, total RNAs were harvested from WT and *Prrc2b*^tm1b-/-^ mice. Another step was added to prevent potential contamination of the isolated RNA with genomic DNA. To do so, the RNA pellet was treated with DNase I (NEB) and incubated for 10 minutes at 37°C. Next, 50 mM EDTA was added to the digested RNA, and the mixture was incubated at 65°C for 5 minutes to stop the digestion. Repurification was performed with 100 μL of phenol/chloroform/isoamyl alcohol (25:24:1). Next, the upper layer was transferred to a fresh tube, followed by adding 10 μL of 3 M sodium acetate, 2 μL of glycogen, and 250 μL of absolute ethanol to allow RNA precipitation. Lastly, the pellet was washed once with 75% ethanol, then centrifuged again to obtain the RNA pellet. PolyA enrichment was performed before library construction by NGS Library Prep. Paired-end sequencing was conducted at Novogene using NovaSeq 6000 S4 with a depth of 20 million reads/sample. Reads were demultiplexed using bcl2fastq version 2.19.0. Quality filtering and adapter removal were performed using Trimmomatic version 0.36 (42) with the following parameters: "ILLUMINACLIP:2:30:10 LEADING:3 TRAILING:3 SLIDINGWINDOW:5:25 MINLEN:32 HEADCROP:10". Processed/cleaned reads were then mapped to the Homo sapien reference genome (GRCh38, hg38) with Hisat version 2.1.1 (43) with the default settings. The subread-2.0.1(44) package (featureCounts) was used to derive gene counts using the gencode.v38 gene annotations (45) with the following parameters: “-T 10 -g gene_name -B -C -p --ignoreDup --fracOverlap 0.1”. DESeq2 version 1.38.3 (46) was used within an R-4.2.2 (47) (URL: https://www.R-project.org/) environment to normalize raw counts and identify significantly changed genes. Gene ontology analyses were performed on significantly changed genes using the clusterProfiler package version 4.6 (48).

### Single-nucleus RNA-sequencing (snRNA-seq) analysis

Single-nucleus RNA sequencing (snRNA-seq) was performed by SingulOmics Corporation using their established protocols. Whole hearts were harvested from both WT and *Prrc2b*^tm1b-/-^ mice, which were at an age of E18.5 (N = 3), and were fast frozen in liquid nitrogen. Nuclei were isolated from frozen mouse heart tissue using the 10x Genomics Single Nuclei Isolation Kit, following the manufacturer’s instructions. The snRNA-seq libraries were constructed using the 10x Genomics Chromium System and the 10x Chromium Multiome kit. Each library was sequenced on the Illumina NovaSeq 6000 platform, generating approximately 200 million paired-end reads (PE150) per library. snRNA-seq reads were demultiplexed and aligned to the mouse reference genome mm10 using 10x Genomics Cell Ranger ARC 2.0.2 (49). A count matrix was created by summarizing reads mapped to both exon and intron regions of each gene in each nucleus.

### Dimension reduction, clustering, annotation, and differential expression analysis for snRNA-seq

Only nuclei with more than 200 snRNA-seq reads fragments were included in the analysis. Potential doublets were eliminated using DoubletFinder (50), with an estimated doublet rate of 5%. Dimension reduction and clustering primarily relied on the top 3000 variant features in snRNA-seq data. Before these processes, data from both WT and KO hearts were integrated, utilizing SCTransformed (51) counts and relevant functions in Seruat v4 (52) in an R-4.2.2 environment. Dimension reduction was initiated with principal component analysis (PCA), followed by uniform manifold approximation and projection (UMAP) analysis using the first 20 PCs. Clustering was performed using the k-nearest neighbors and Shared Nearest Neighbor (SNN) algorithms (53). Cluster annotations were initially established by comparing gene expression profiles with publicly available annotated datasets using SingleR (54), with gene expression counts normalized and scaled prior to these comparisons. Reference datasets from Tabula Muris (55), containing snRNA-seq data from mouse heart and aorta, were employed for this annotation, and additional refinement was undertaken by considering normalized RNA expression and gene accessibility data for well-established cardiac cell-type-specific markers (56) (57). To identify differentially expressed genes (DEGs) within each cluster, a logistic regression model, known for its robust performance in previous work (58) (59), was applied to normalized counts, with variations in sequencing depth across nuclei treated as latent variables. The DEG analysis included only genes expressed in more than 10% of the cluster. Bonferroni correction was applied to adjust the nominal *P* values; genes with |Log2 fold change| > 0.2 and adjusted *P* < 0.05 were considered significant.

### Zebrafish maintenance and microinjection of morpholinos

Adult and larval zebrafish husbandry and care were conducted in full accordance with animal care and use guidelines, with ethical approval by the University Committee on Animal Resources at the University of Rochester Medical Center. Adult wild-type Tübingen zebrafish were maintained on a 14h:10 h (light: dark) cycle, and newly fertilized embryos were collected for morpholino (MO) injection and phenotypic characterization. Morpholinos were designed against the 5’-terminal sequence near the start codon (ATG) and the junction of exon 3 intron 3 (e3i3) of the *Prrc2b* gene to block mRNA translation and trigger exon 3 exclusion or intron retention, respectively. *Prrc2b* MO (Translation-blocking Morpholino): 5’- ATTTGCCCCAAACGATCGGACATTG -3’; *Prrc2b* MO (e3i3 Splicing-altering Morpholino): 5’- TACAACACACACGCCGTACCTCC -3’; and a standard negative control MO (5’- CCTCTTACCTCAGTTACAATTTATA -3’) were injected into one to two cell stages of zebrafish embryos. Images of zebrafish larvae were taken using a dissecting microscope.

### Quantification and statistical analysis

The data were analyzed using GraphPad Prism V9.3.1 and expressed as mean *±* standard deviation (SD) as described in the figure legends. Statistical analyses between two groups were performed using an unpaired Student’s t-test, while a one-way, two-way, or three-way analysis of variance (ANOVA) test was used for cases involving more than two groups with two or more independent variables. Statistical significance was considered when *P* < 0.05.

## Data Availability

All data produced in the present study are available upon reasonable request to the authors.

## Acknowledgments

We thank Jiangbin Wu, Jared Hollinger, Emily Bonanno, and Matthew Auguste for their technical assistance and Tianlong Zhang for his help with structure prediction using the AlphaFold web tool. We also thank Belal Chowdhury for reading this manuscript and providing critical scientific comments to improve it. We appreciate the technical assistance from Erika Flores Medina in histology, Amy Mohan, Deanne Mickelsen, and Christine Christie in surgical operations (Aab CVRI), and the Genomic Research Center for the bulk next-generation RNA-sequencing analysis. We thank INFRAFRONTIER/EMMA for providing the mutant mouse line (C57BL/6N- A<TM1BRD>*Prrc2b*<tm1a(EUCOMM)Wtsi>/WtsiBiat (frozen sperms)), INFRAFRONTIER/EMMA (www.infrafrontier.eu, PMID: 25414328), from which the mouse line was distributed (EM:05981). Associated primary phenotypic information of the *Prrc2b* tm1a mice may be found at the International Mouse Phenotyping Consortium. We also appreciate the technical assistance from Lin Gan for *in vitro* fertilization and for generating the *Prrc2b* tm1a mouse line on a service fee basis, and from Lynne Maquat for using their polysome fractionator. We acknowledge the Pediatric Cardiac Genetics Consortium (PCGC) for providing published data on genetic variants in the PRRC2B gene from 3,740 CHD probands in the PCGC genetic mutation databases.

## Author contributions

PY launched the study and obtained the funding. PY, DD, AVI, ESK, and FJ conceived the ideas, designed the experiments, analyzed the data, and wrote the manuscript. DD, AVI, ESK, FJ, LY, YK, LW, HL, and FM conducted the experimental work. FJ, SZ, and JG performed the bioinformatic analysis. GAP, ZJ, PM, and JG provided technical assistance, conceptual feedback, and contributed to the writing and editing of the manuscript. All the authors discussed the results and had the opportunity to comment on the manuscript.

---

**Figure S1.**
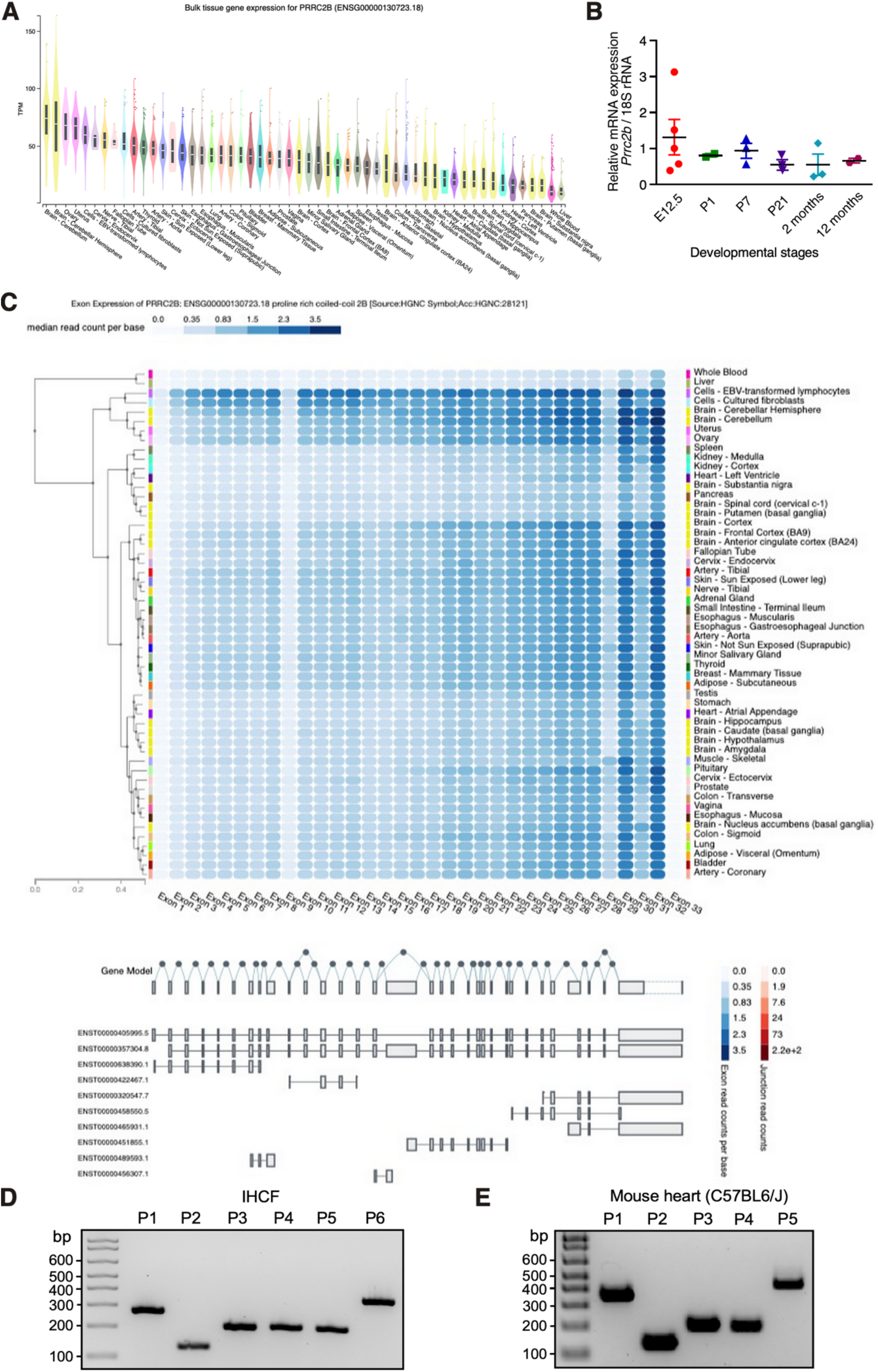
Alternative splicing patterns of *PRRC2B* in human organs. **A**. Bulk tissue gene expression for *PRRC2B* mRNA in human organs from the GTEx Portal databases. **B**. *Prrc2b* mRNA expression in mouse hearts at different developmental stages. **C**. Alternative splicing isoforms of *PRRC2B* mRNA across different human organs. **D**. RT-PCR validation of alternative splicing isoforms of *PRRC2B* mRNA in IHCF cells. P1-6, primers 1-6. **E**. RT-PCR validation of alternative splicing isoforms of *Prrc2b* mRNA in the mouse heart (from a 2- month-old C57BL6/J male mouse). P1-5, primers 1-5.

**Figure S2.**
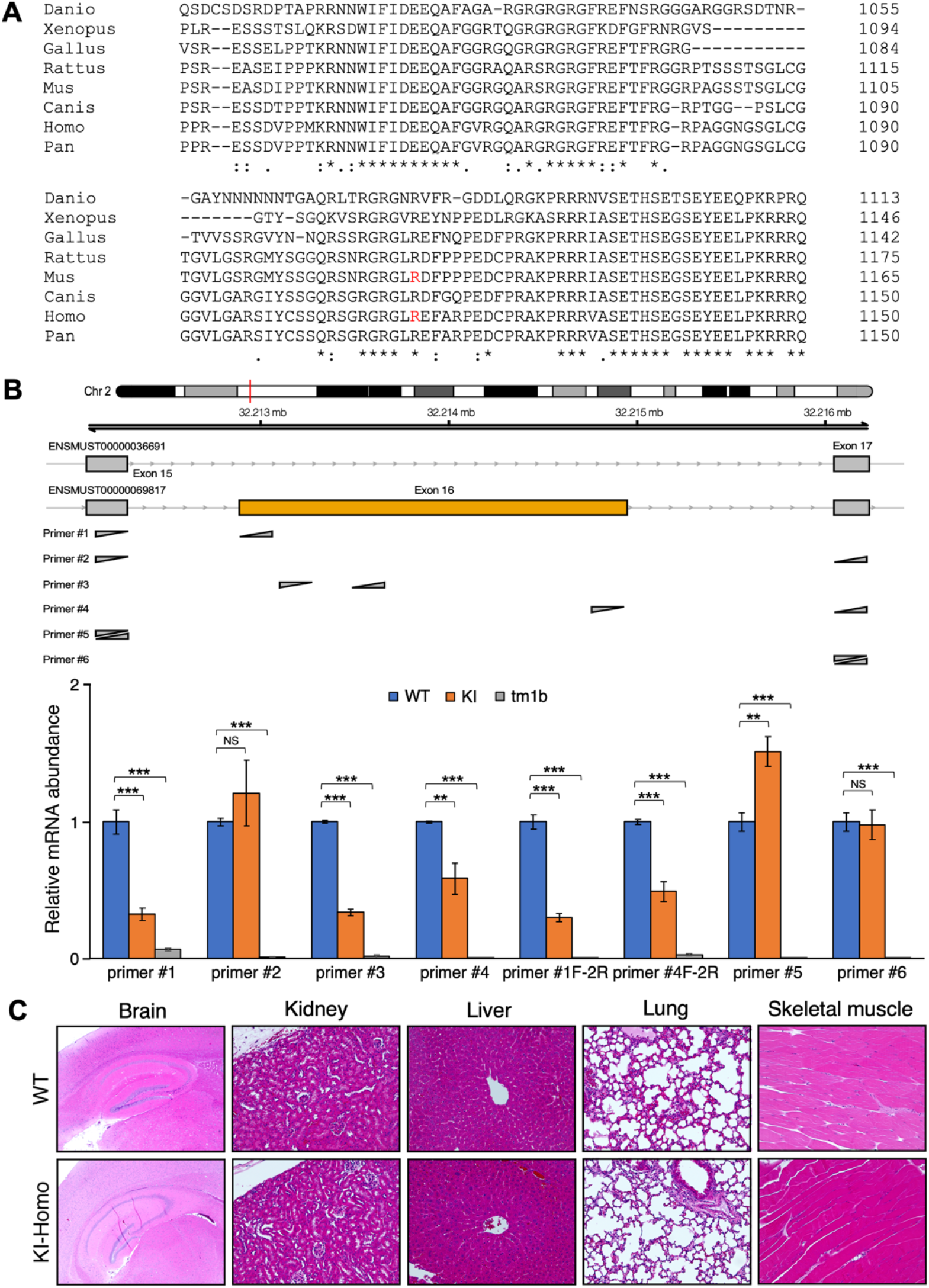
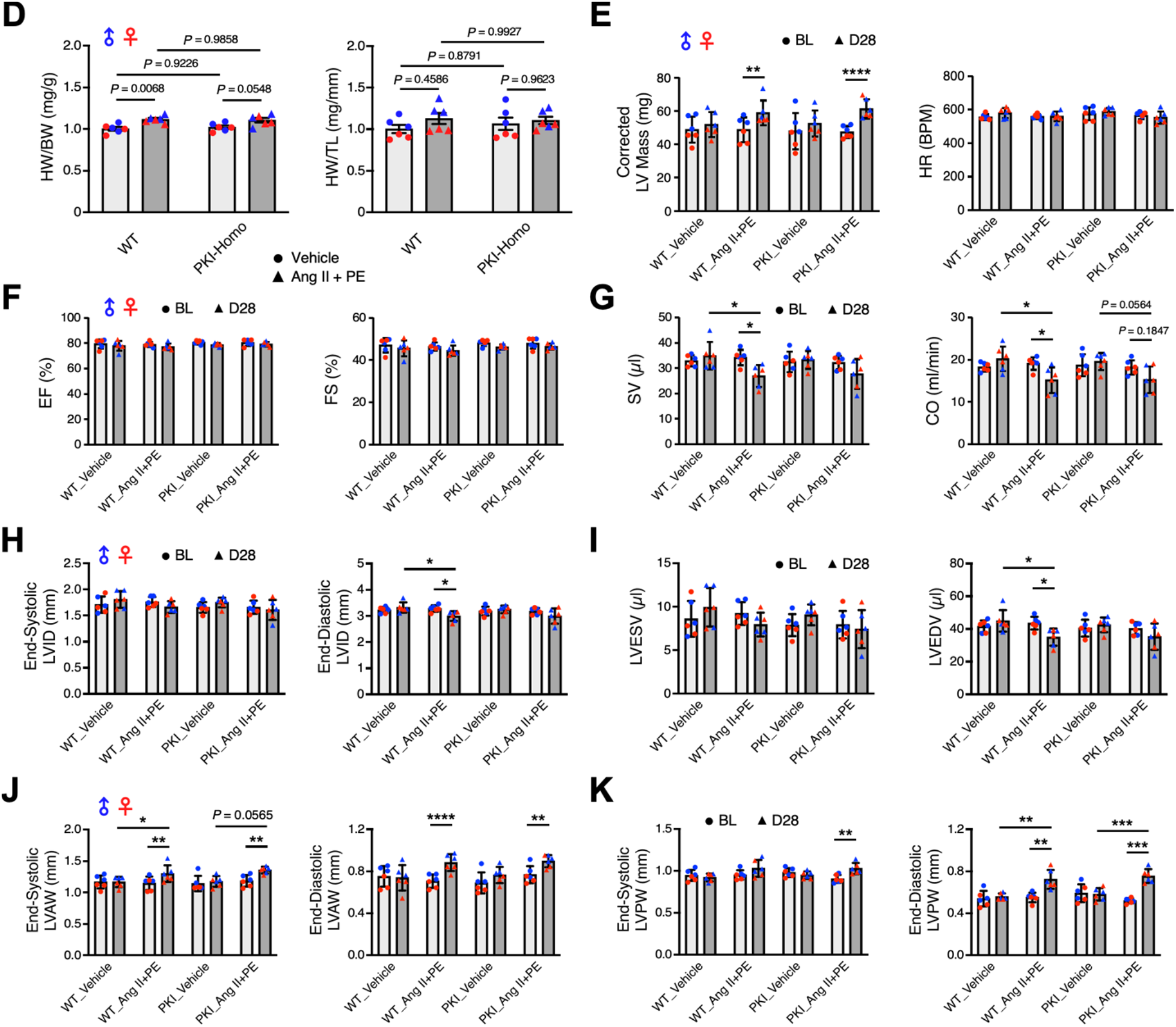
Characterization of *Prrc2b*^R1128X/R1128X^ global knock-in mice. **A.** Evolutionary conservation of the PRRC2B protein sequence surrounding the R1113 amino acid residue in humans across various species (R1128 in mice). **B**. RT-qPCR detection of alternative splicing isoforms of *Prrc2b* mRNA in *Prrc2b*^R1128X/R1128X^ global knock-in (FL-*Prrc2b*^gKO^) and tm1b (*Prrc2b*^gKO^; KO of both FL and ΔE16 alternative spliced isoform of *Prrc2b* mRNAs) mice. **C**. Representative H&E images of multiple organs of WT and *Prrc2b*^R1128X/R1128X^ global knock-in mice. **D-K**. Quantitative comparisons of the morphological and echocardiographic difference between the *Prrc2b*^gKO^ and WT mice, including heart weight (HW)/body weight (BW) and heart weight (HW)/tibia length (TL) (**D**), corrected left ventricle (LV) mass and heart rate (HR) (**E**), ejection fraction (EF) and fractional shortening (FA) (**F**), stroke volume (SV) and cardiac output (CO) (**G**), end-systolic LV internal diameter and end-diastolic LV internal diameter (LVID) (**H**), LV end-systolic volume (ESV) and end-diastolic volume (EDV) (**I**), end-systolic LV anterior wall and end-diastolic LV anterior wall (LVAW) thickness (**J**), and end-systolic LV posterior wall and end- diastolic LV posterior wall (LPAW) thickness (**K**). Data are represented as mean ± SD. A three- way ANOVA was performed to compare the four groups with repeated measures on time, followed by Tukey’s comparisons in E-K (mean ± SEM in D, analyzed by two-way ANOVA followed by Tukey’s multiple comparisons). * *P* < 0.05; ** *P* < 0.01; *** *P* < 0.001; **** *P* < 0.0001.

**Figure S3.**
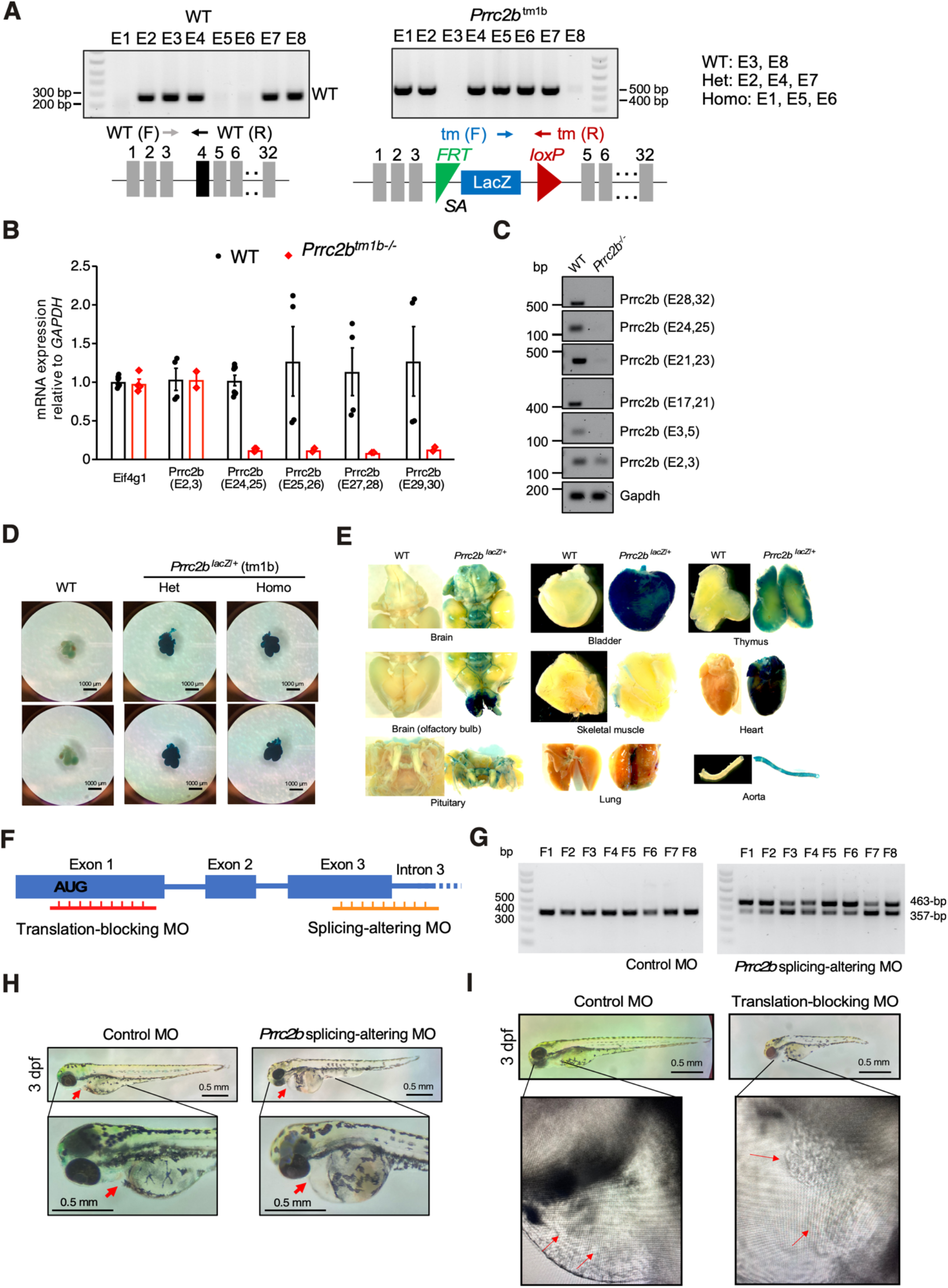
Generation of *Prrc2b*^tm1b-/-^ global knockout mice (*Prrc2b*^-/-^) and cardiac developmental defects in zebrafish upon ASO-mediated *Prrc2b* inactivation. A. Representative genotypes of *Prrc2b*^-/-^, *Prrc2b*^+/-^, and *Prrc2b*^WT^ at E13.5 determined by two pairs of primers (6tm, F and 7tm, R) and (WT, F and WT, R). **B**. RT-qPCR measurement of *Prrc2b* mRNA expression normalized by *Gapdh* mRNA. **C**. Representative agarose gel electrophoresis of PCR products from cDNA using two pairs of primers (Ex2, F and Ex3, R), (Ex3, F and Ex5, R), (Ex17, F and Ex21, R), (Ex21, F and Ex23, R), (Ex24, F and Ex25, R) & (Ex28, F and Ex32, R) and normalized with *Gapdh*. **D**. LacZ staining of embryonic hearts at E12.5. **E**. LacZ staining of multiple organs in WT and *Prrc2b*^+/-^ adult mice. **F**. Schematic of *Prrc2b*-inactivating antisense morpholino oligomers (MO). Translation-blocking MO inhibits the translation of *Prrc2b* mRNA. Splicing-altering ASOs either skip exon 3 or include intron 3, and the latter outcome was observed. **G**. Representative agarose gel electrophoresis images of PCR products from cDNA upon RT of total RNA harvested from control MO- and *Prrc2b* splicing-altering MO-injected embryos determined by primer pair (e1E2, F & E4e5, R). **H**. *Prrc2b* splice-exon-3 MO was injected into 1–4 cell stage in zebrafish embryos to cause aberrant *Prrc2b* mRNA splicing and nonsense- mediated mRNA decay. A total of five *Prrc2b* splicing-altering MO-injected embryos showed curved bodies with pericardial edema. Representative images were shown. Scale bar: 0.5 mm. **I**. *Prrc2b* translation-blocking MO was injected into 1–4 cell stage in zebrafish embryos to inhibit *Prrc2b* mRNA translation. A total of four translation-blocking MO-injected embryos showed curved bodies with pericardial edema. Representative images were shown. Scale bar: 0.5 mm.

**Figure S4.**
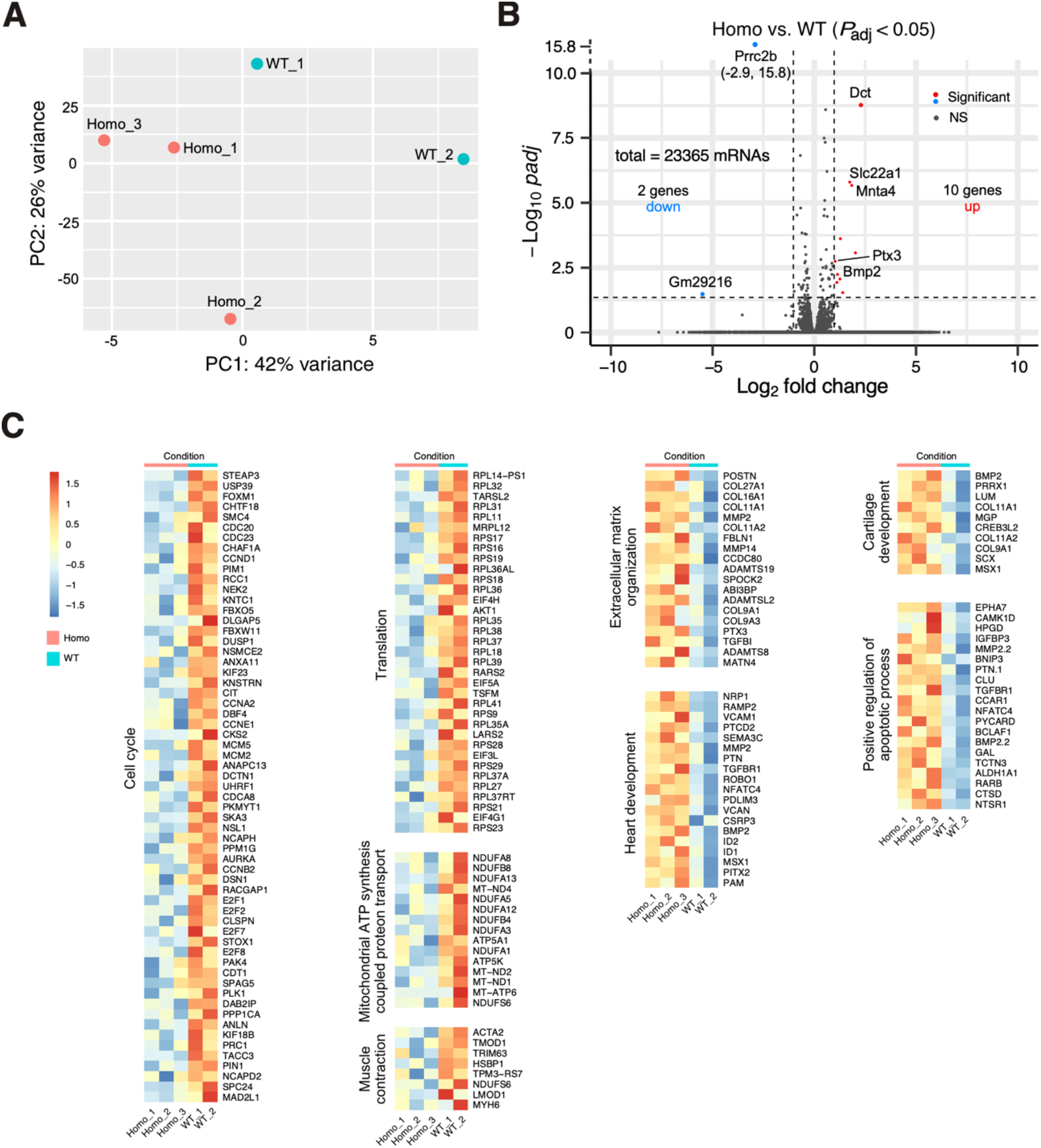
RNA-seq analysis of *Prrc2b* global knockout hearts at E18.5. **A**. Principal component analysis (PCA) of RNA-seq data for WT and Homo *Prrc2b*^tm1b-/-^ KO hearts at E18.5. **B**. Volcano plot of RNA-seq of WT and *Prrc2b*^tm1b-/-^ hearts. Log_2_ Fold Change (*Prrc2b*^tm1b-/-^ / WT) is plotted as the X-axis, while -Log10 *P*_adj_-values are plotted as the Y-axis. Genes with |Log2 Fold Change| > 1 and *P*_adj_-value < 0.05 are colored red. **C**. Heatmap of differential gene expression for multiple GO pathways.

**Figure S5.**
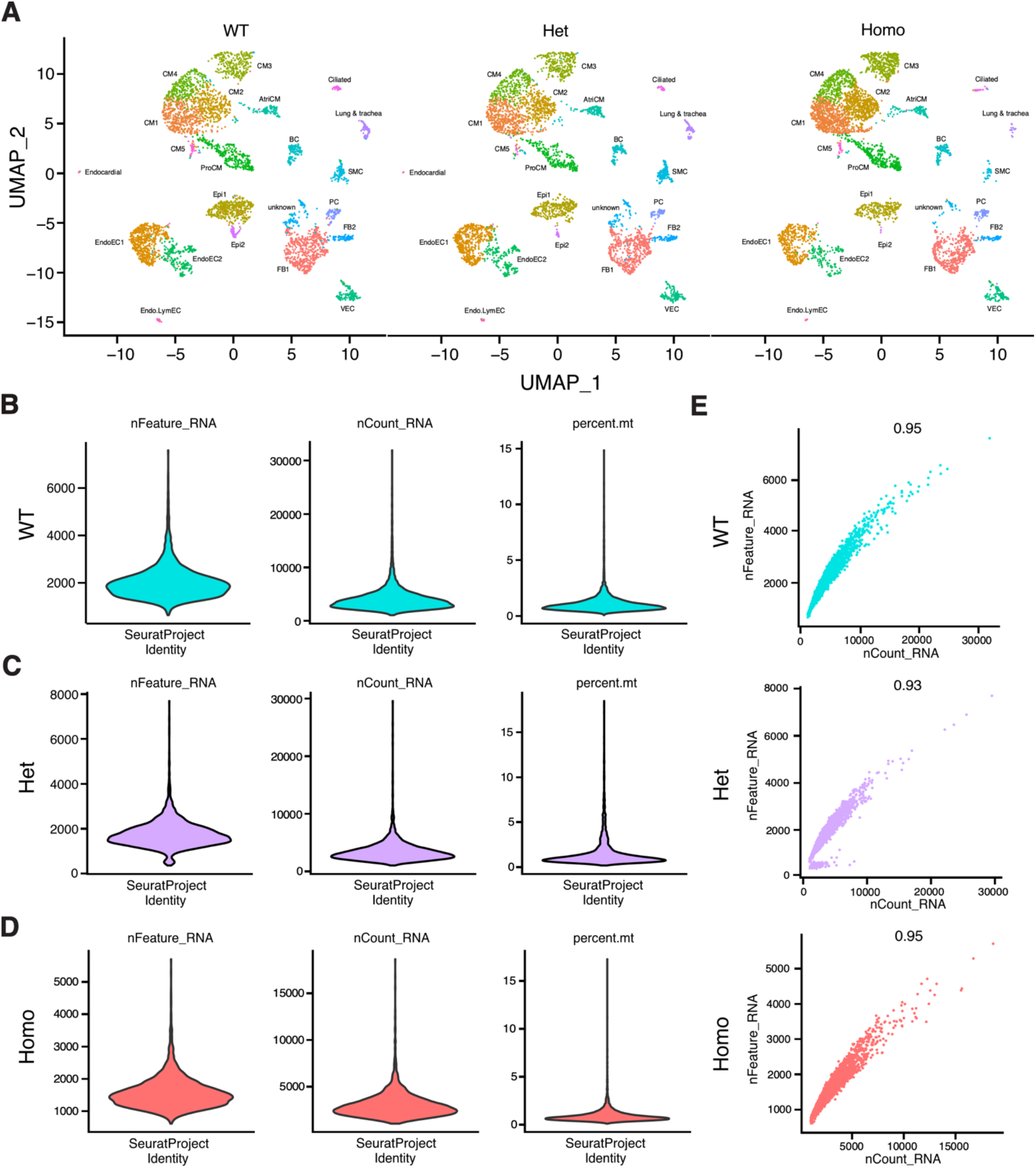
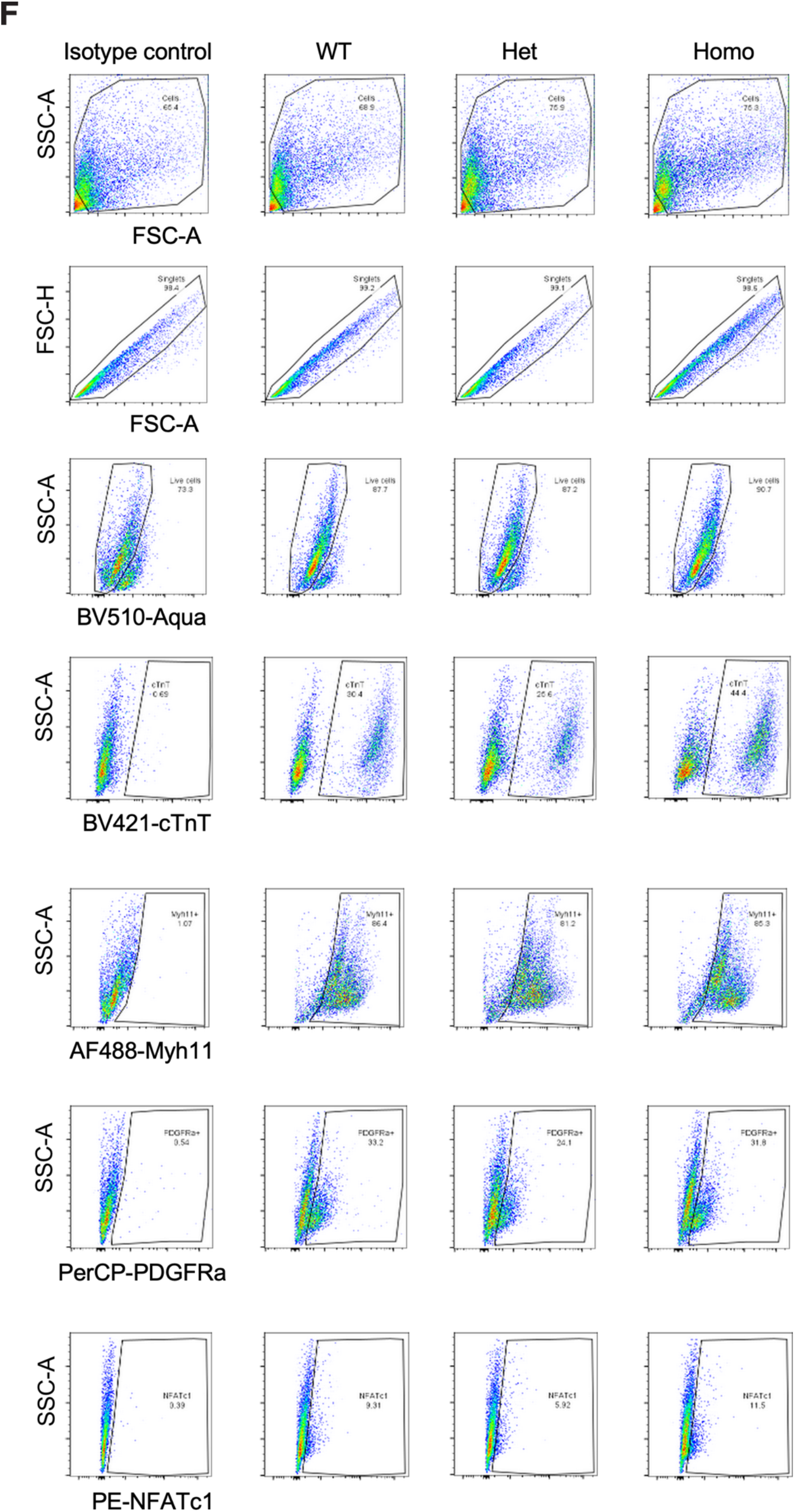
snRNA-seq analysis of *Prrc2b* global knockout hearts at E18.5. **A**. A UMAP (uniform manifold approximation and projection) presentation of clustering results based on the top 3000 variant features. 19 clusters were identified and labeled with cell type and top marker genes. Nuclei from WT, Het, and Homo were clustered together and plotted separately. Ven.CM, ventricular cardiomyocytes; Atri.CM, atrial cardiomyocytes; FB, fibroblasts; EC, endothelial cells; SMC, smooth muscle cells; BC, blood cells; VEC, vascular EC; Epi, epicardial cells; PC, pericytes; ProCM, proliferating CM; Endo, endocardial; Lym, lymphatic. **B-D**. Quality control vlnplots showing the distribution of number of genes (nfeature) (**B**), number of reads (ncount) (**C**), percentage of mitochondrial coded genes (percentage.mt) (**D**) detected in each nucleus in each mouse heart (WT, Het, Homo). **E**. Quality control scatterplots showing the number of genes (nfeature, y-axis) and the number of reads (ncount, x-axis) detected in each nucleus in each sample. The Pearson correlation coefficient is labeled. **F**. Flow cytometry gating strategy used for cardiomyocytes (cTnT+), smooth muscle cells (MYH11+), epicardial cells (NFATc1+), and cardiac fibroblasts (PDGFRα+) in WT, Het, and Homo mice, respectively.

**Figure S6.**
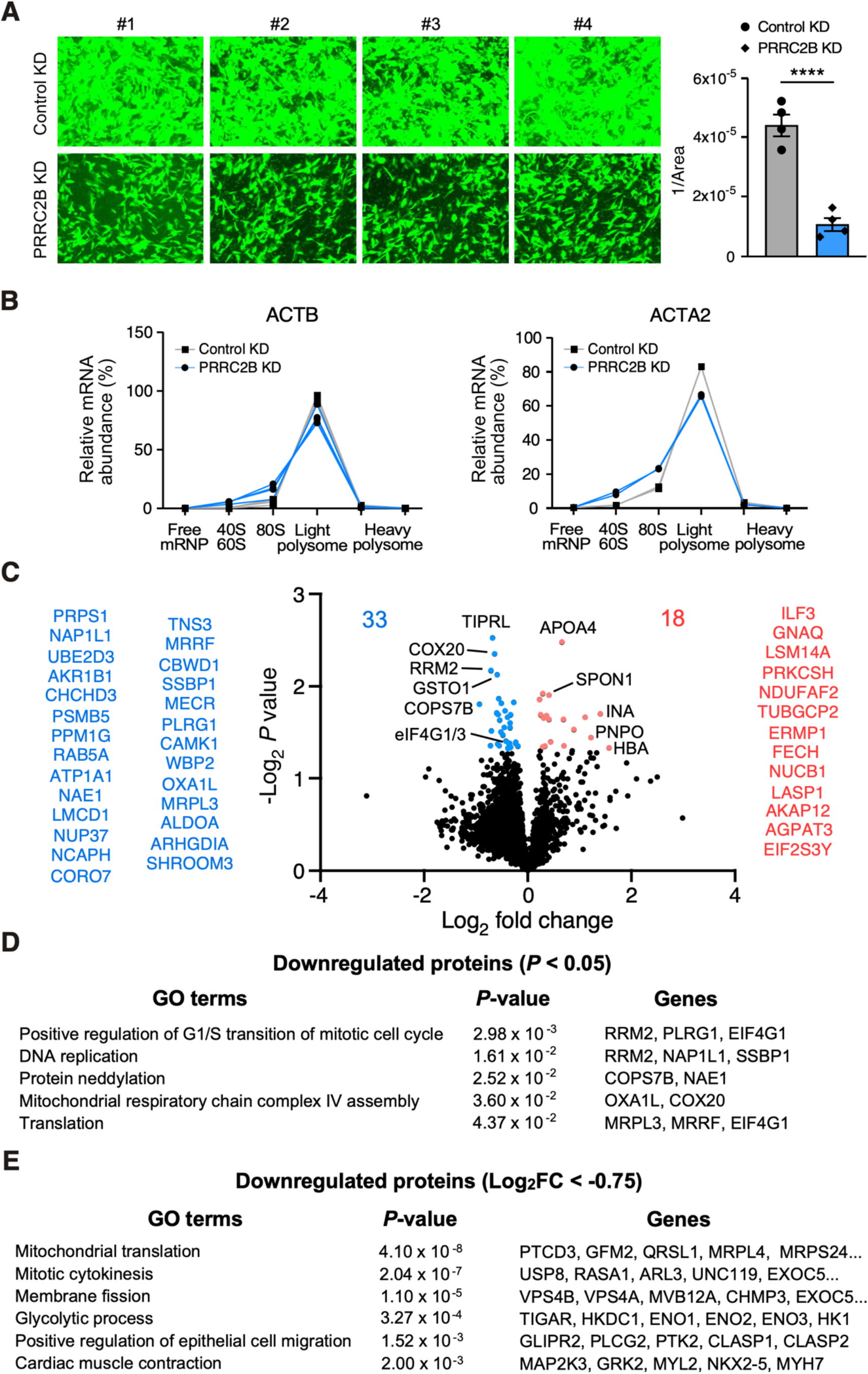
Mass spectrometry analysis of *Prrc2b* global knockout hearts. **A**. Quantitative analyses of cell migration rate using transwell migration assay upon *PRRC2B* knockdown in human aorta-derived smooth muscle cells. **B**. Polysome profiling coupled with RT- qPCR for *ACTB* and *ACTA2* mRNAs in human aorta-derived SMCs upon *PRRC2B* knockdown. This experiment was repeated 2-4 times (as biological replicates), and all the results were shown. **C**. Volcano plot of differentially expressed proteins identified by mass spectrometry in *Prrc2b*^tm1b-/-^ gKO compared with WT control hearts. 33 downregulated and 18 upregulated proteins with statistical significance (*P* < 0.05) were listed. N = 3 E18.5 mouse hearts were used from WT and gKO mice. **D**. Gene ontology analysis of significantly downregulated proteins (*P* < 0.05). **E**. Gene ontology analysis of drastically downregulated proteins (Log_2_FC < -0.75).

**Figure S7.**
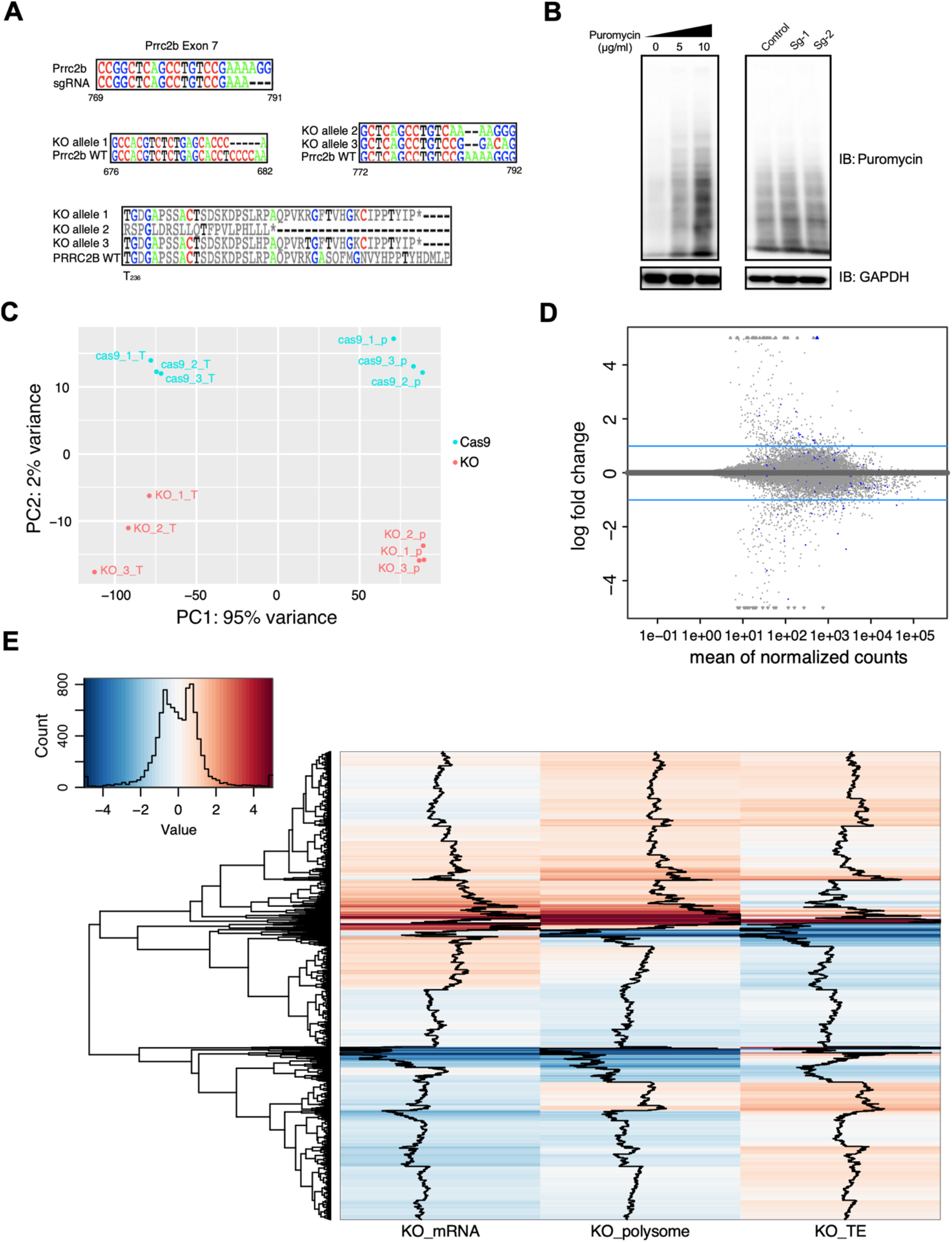
Transcriptomic and translatomic profiling of *PRRC2B* knockout human cells. **A**. Sanger-sequencing of the DNA locus of three copies of the *PRRC2B* gene in knockout HEK293T cells. **B**. Puromycin incorporation assay of control (transfected with Cas9 without guide (g)RNA) and KO HEK293T cells. Sg1 and Sg2 are two single gRNAs for KO. **C**. Principal component analysis (PCA) showing the distribution of the biological triplicates of RNA-seq (T) and polysome-seq (p) for the control and KO cells. **D**. An M (log ratio) versus A (mean average) plot (MA-plot) showing the distribution of differentially expressed genes. Genes with significant *P* values (*P* < 0.05) are colored in blue. **E**. Heatmaps for RNA-seq and polysome-seq, and calculated TE (translation efficiency) showing the annotated dysregulated genes in all samples. The black lines indicate the average fold change per gene across the dataset.

**Figure S8.**
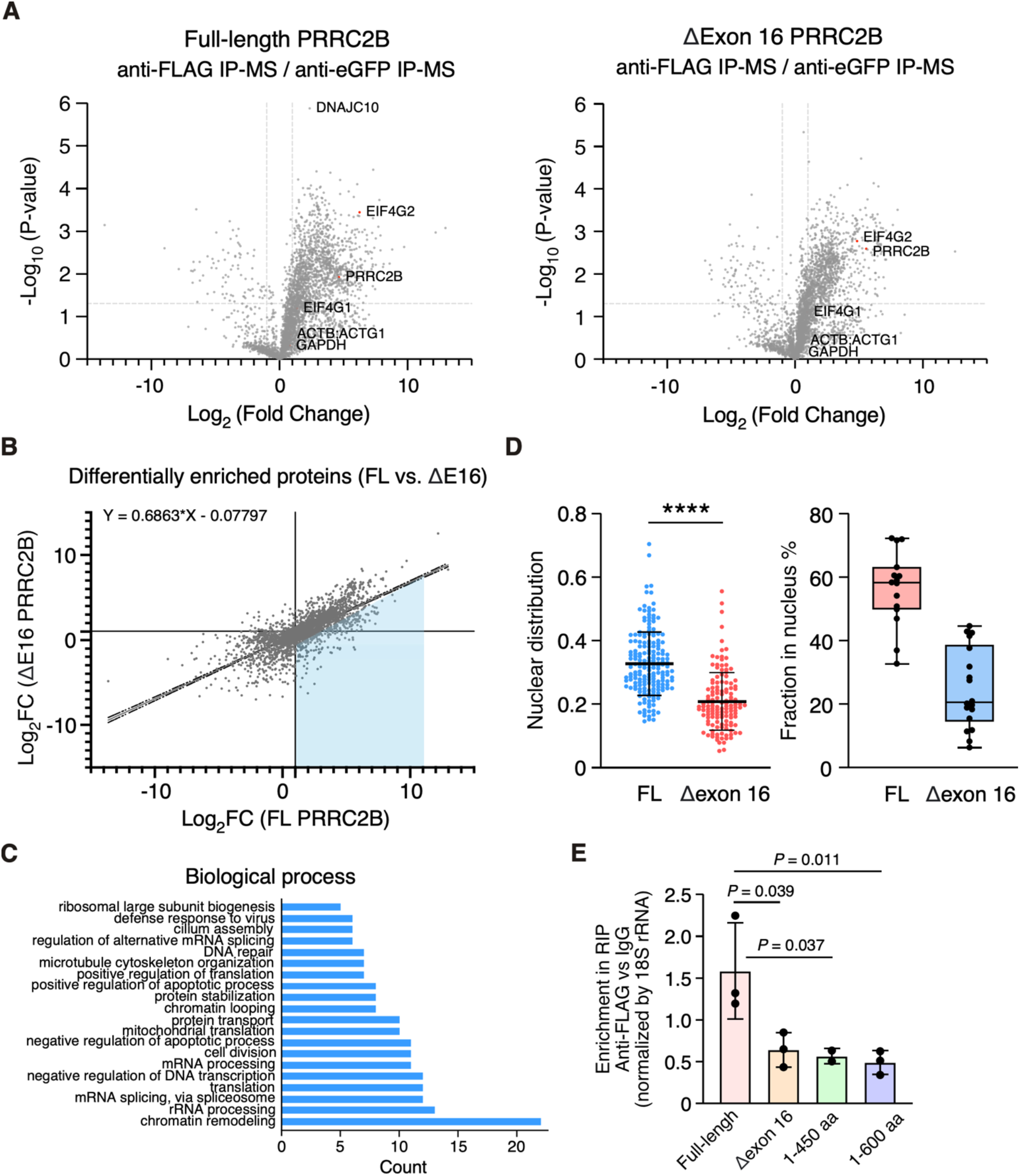
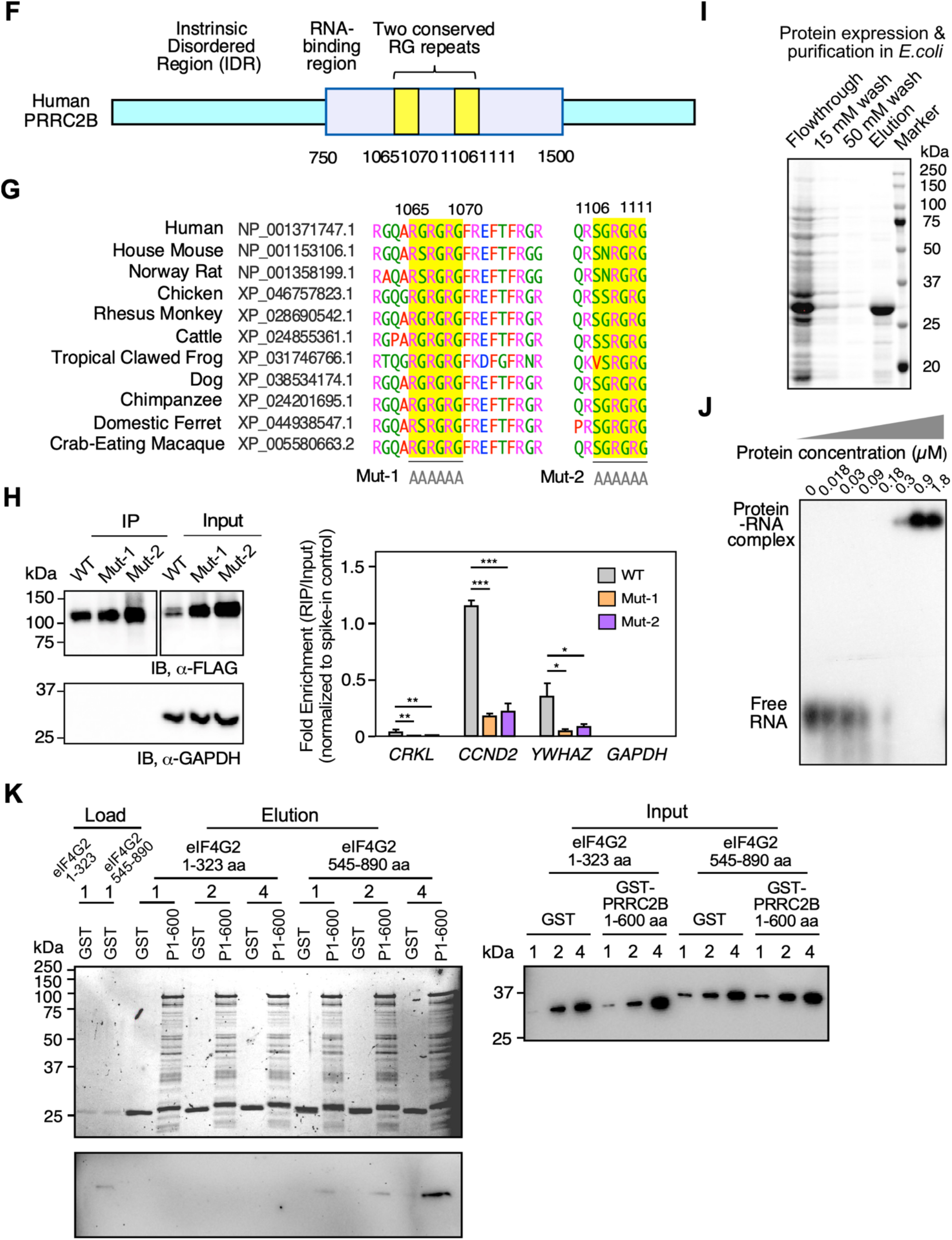
Interactome of alternative splicing isoforms of PRRC2B and interacting domain mapping of PRRC2B-eIF4G2 complex and PRRC2B-RNA binding. **A**. Immunoprecipitation (RNase A-treated) and mass spectrometric identification of PRRC2B protein interactome for both full-length and ΔE16 alternative spliced protein isoforms after overexpression in HEK293T cells. **B**. Differentially enriched interacting proteins comparing full- length and ΔE16 PRRC2B alternative spliced protein isoforms. **C**. Gene ontology analysis of unique binding protein partners of full-length PRRC2B protein isoform showing enriched interacting proteins in the nucleus compared to the ΔE16 alternative spliced isoform. **D**. Quantification of nuclear localization of full-length and ΔE16 alternative spliced isoforms after overexpression regarding the signal intensity and percentage of cells in HEK293T cells. **E**. RIP- RT-qPCR for *CCND2* mRNA for full-length PRRC2B, ΔE16 alternative spliced isoform, and N- terminal domains of PRRC2B (1-450 aa and 1-600 aa) upon overexpression in HEK293T cells. **F**. Schematic model of two key ArgGly (RG)-rich RNA-binding motifs in full-length PRRC2B protein. **G**. Evolutionary conservation of the key RG-rich RNA-binding motifs across vertebrates, including frogs. **H**. Mutagenesis of RNA-binding motifs of PRRC2B-3xFLAG. RIP-RT-qPCR was performed after IP using FLAG antibody to detect well-established PRRC2B-bound target mRNAs (i.e., *CCND2*, *YWHAZ*, and *CRKL*) for full-length WT and mutant variants of PRRC2B (Mut-1 and Mut-2). **I**. Expression and purification of truncated PRRC2B protein (900-1200 aa) in BL21(DE3) with imidazole buffer wash. **J**. Electrophoresis mobility shift assay (EMSA) of 900-1200 aa PRRC2B fragment with *CCND2* mRNA 5’-UTR (100-nt) in a binding buffer. **K**. Domain mapping for the interacting interface between PRRC2B and eIF4G2 using mutagenesis, in-cell overexpression, and IP-IB. Data are represented as mean ± SD. An unpaired two-tailed Student t-test was performed to compare two groups for D. A Lognormal One-way ANOVA with Tukey post-hoc test was performed to compare the four groups in E and the three groups in H. * *P* < 0.05; ** *P* < 0.01; *** *P* < 0.001; **** *P* < 0.0001.

**Table S1.** PRRC2B variant burden test in congenital heart disease (CHD) probands

| Phenotype (N) | Gene Wide Burden | Variants at RNA binding sites | Variants at arginine sites | Gene Wide Burden: Ultrarare |
| --- | --- | --- | --- | --- |
| **LVOTO (863)** | 0.14133 | 0.08867 | 0.28310 | 0.31683 |
| **HLHS (399)** | 0.02508 | 0.84289 | 0.62052 | 0.14477 |
| **Heterotzxy (345)** | 0.93369 | 0.94747 | 0.84024 | 0.64972 |
| **ASD (167)** | 0.00002 | 0.98771 | 0.00001 | 0.04726 |
| **ToF (702)** | 0.87008 | 0.84625 | 0.67232 | 0.40356 |
| **Conotruncal (2569)** | 0.22075 | 0.19740 | 0.04527 | 0.46164 |

**Table S2.**
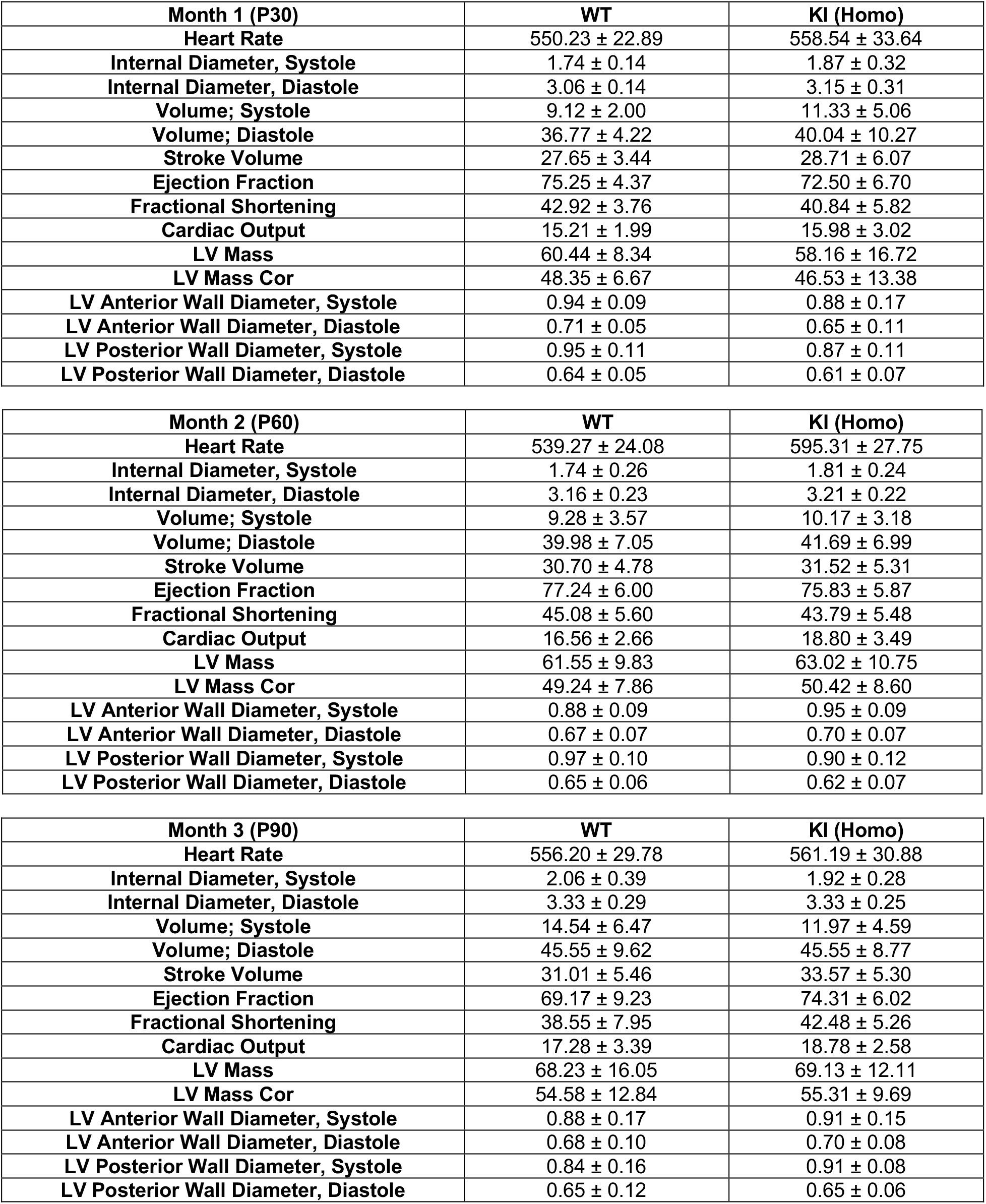
Echocardiography of WT and *Prrc2b*^p.R1128X/p.R1128X^ KI (FL-*Prrc2b*^gKO^) mice at the age of 1-3 months at baseline. Data is shown as mean ± SD. Technical replicates for each biological sample are plotted (WT: N = 13; KI: N = 14). A two-tailed, unpaired Student t-test was performed to compare two groups. * *P* < 0.05; ** *P* < 0.01.

**Table S3.** Differentially expressed genes in the whole hearts from the bulk RNA-seq of WT and *Prrc2b*^tm1b-/-^ mice at E18.5 (N = 3 WT and N = 2 *Prrc2b*^tm1b-/-^ from the same litter to avoid batch effects).

**Table S4.** Differentially expressed genes in smooth muscle cells and cardiomyocytes discovered by the single nucleus (sn)RNA-seq of WT and *Prrc2b*^tm1b-/-^ mice at E18.5 (N = 3 WT and N = 3 *Prrc2b*^tm1b-/-^ hearts combined).

**Table S5.** Differentially translated mRNAs in whole hearts revealed by polysome-seq analysis normalized by RNA-seq of WT and *Prrc2b*^tm1b-/-^ mice at E18.5 (N = 3 WT and N = 3 *Prrc2b*^tm1b-/-^ hearts separately).

**Table S6.** Differentially expressed proteins in whole hearts revealed by the quantitative mass spectrometry analysis of WT and *Prrc2b*^tm1b-/-^ mice at E18.5 (N = 3 WT and N = 3 *Prrc2b*^tm1b-/-^ hearts separately).

**Table S7.** Differentially expressed genes (RNA-seq) and differentially translated mRNAs (polysome-seq normalized by RNA-seq) in WT control (mock edited) and *Prrc2b* KO HEK293T cells (N = 3 WT and N = 3 KO cells as biological replicates).

**Table S8.** IP-mass spectrometry analysis using anti-FLAG or pre-immune IgG in HEK293T cells with overexpression of FLAG-tagged FL or ΔE16 PRRC2B in the presence of RNase A treatment **(N = 3 biological replicates)**.

**Table S9.**
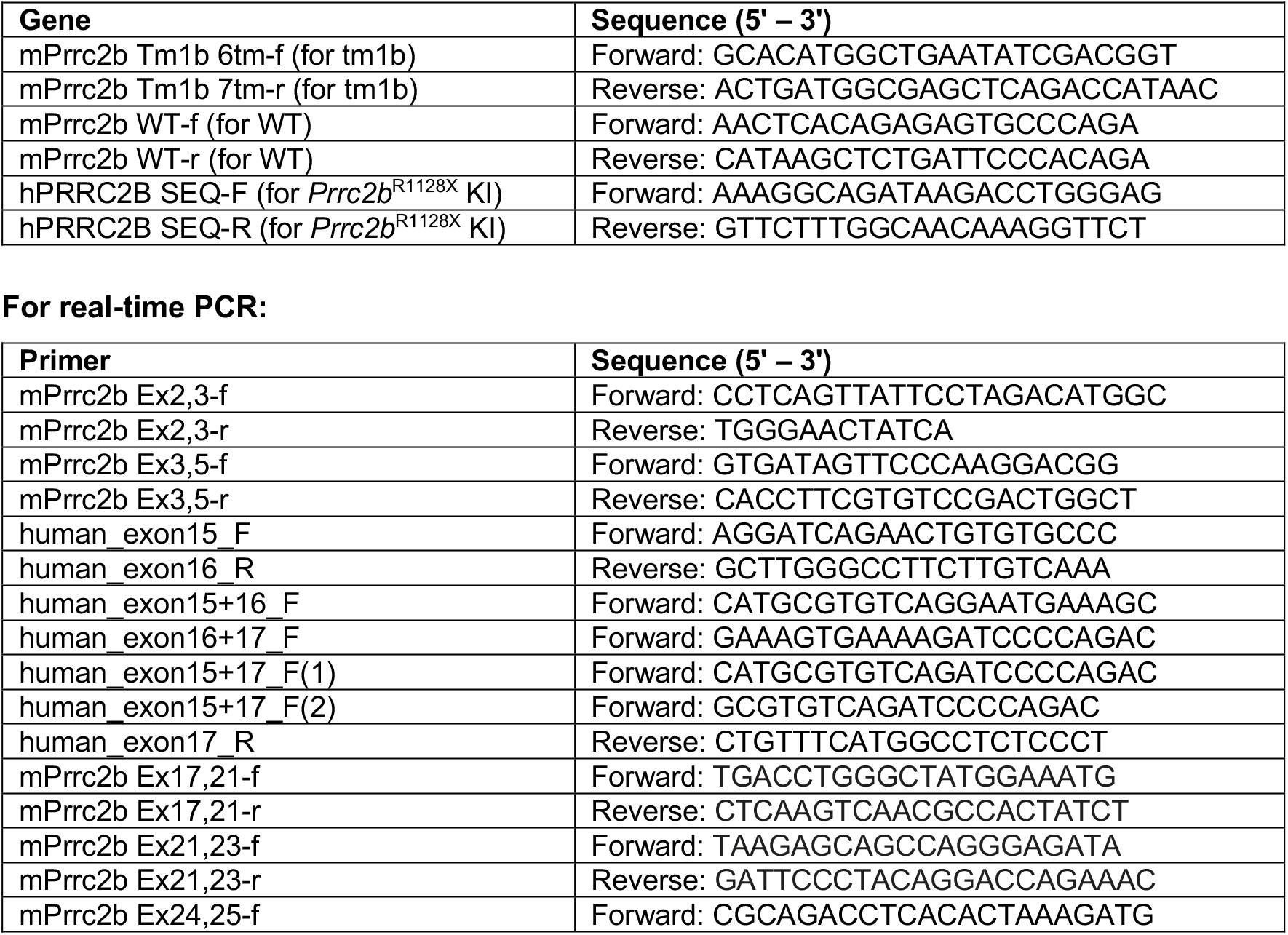
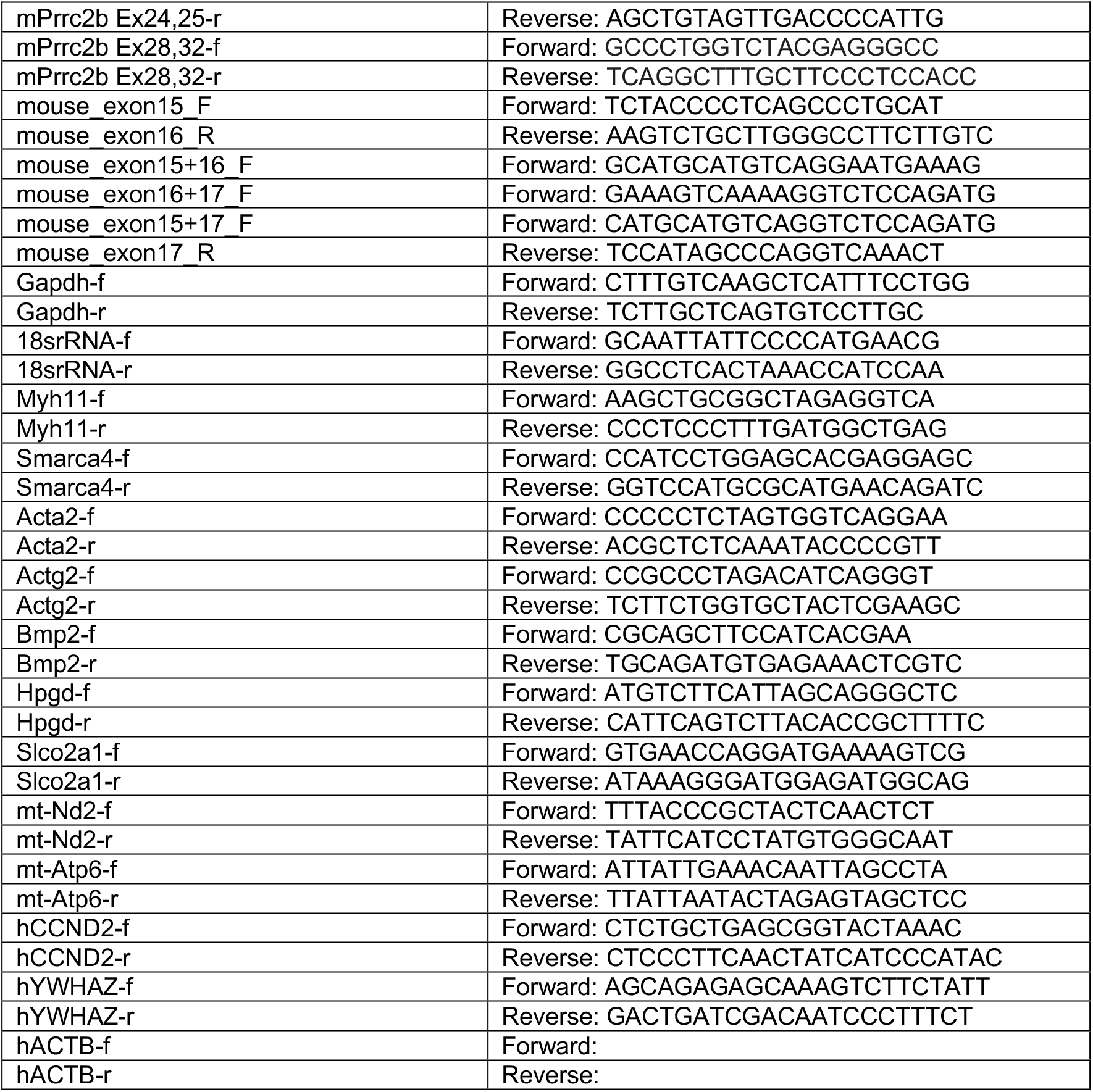
List of primer sets used in this study.

**Table S10.** List of antibodies used in this study

| Antibody | Vendor & Catalogue number |
| --- | --- |
| Anti-eIF4G2 | Proteintech, Cat. #17728-1-AP |
| Anti-PRRC2B | ThermoFisher, Cat. #PA5-66677 |
| anti-GAPDH | Proteintech, Cat. #60004-1-Ig |
| Anti-α-tubulin | Proteintech, Cat. #66031-1-1g |
| Anti-6xHis | Proteintech, Cat. #6605-1-1g |

**Antibodies used for Immunoprecipitation (IP):**
| Antibody | Vendor & Catalogue number |
| --- | --- |
| Anti-eIF4G2 | Proteintech, Cat. #17728-1-AP |
| Anti-FLAG | Proteintech, Cat. #66008-2-Ig |
| Anti-IgG | Cell Signaling Technology, Cat. #2729S |

**Antibodies used for Immunofluorescence (IF):**
| **Antibody** | **Vendor & Catalogue number** |
| --- | --- |
| Anti-eIF4G2 | Proteintech, Cat. #17728-1-AP |
| Anti-FLAG | Proteintech, Cat. #66008-2-Ig |

**Antibodies used for Flow Cytometry (FC):**
| **Antibody** | **Vendor & Catalogue number** |
| --- | --- |
| Anti-cTnT | BD Horizon, Cat. #565618 |
| Anti-MYH11 | Abcam, Cat. #ab320557 |
| Anti-NFATC1 | BioLegend, Cat. #649605 |
| Anti-PDGFRα | eBioscience, Cat. #46-1401-80 |
| BV421 Mouse IgG1 | BD Horizon, Cat. #562438 |
| IgG2a kappa Alexa Fluor 488 | eBioscience, Cat. #53472480 |
| PE Mouse IgG1 | BioLegend, Cat. #400111 |
| PerCP IgG2a kappa (eBR2a) | eBioscience, Cat. #46432180 |

